# Time-dependent relationship between urinary biomarkers of nucleic acid oxidation and colorectal cancer risk

**DOI:** 10.1101/2025.01.21.25320898

**Authors:** Yingya Zhao, Marina S. Nogueira, Ginger L. Milne, Yu-Tang Gao, Qiuyin Cai, Qing Lan, Haoyang Yi, Nathaniel Rothman, Xiao-Ou Shu, Wei Zheng, Qingxia Chen, Gong Yang

## Abstract

**PURPOSE:** Randomized controlled trials have failed to validate that neutralizing oxidative stress (OxS) through antioxidant supplementation reduces cancer risk. This study aims to prospectively investigate whether the relationship between systemic OxS and colorectal cancer (CRC) risk changes over the course of cancer development.

**METHODS:** This study utilized a nested case-control design in two Shanghai cohorts for primary analysis and one US cohort for replication analysis. During a median follow-up of 15.1 years in the Shanghai cohorts, 1938 incident CRC cases were identified and matched to one control each. In the US cohort, 285 incident CRC cases were included with two matched controls per case. Systemic OxS was assessed by urinary markers of DNA oxidation (8-oxo-7,8-dihydro-2’-deoxyguanosine [8-oxo-dG]) and RNA oxidation (7,8-dihydro-8-oxo-guanosine [8-oxo-Guo]) using UPLC-MS/MS assays. Multivariable-adjusted odds ratios (ORs) for CRC risk were calculated.

**RESULTS:** After adjusting for potential confounders, we observed an inversion association between OxS markers and CRC risk in the Shanghai cohorts, which was independently replicated in the US cohort. Moreover, the inverse association was time-dependent, manifesting only for CRC cases diagnosed within 5 years of enrollment. ORs (95% CI) for CRC at the 10th and 90th percentiles of 8-oxo-dG levels, relative to the median, were 1.87 (1.39 to 2.53) and 0.48 (0.37 to 0.63), respectively, demonstrating an approximate 4-fold difference in risk between the two groups, with *P* for overall association of < 0.001. A similar pattern was observed for 8-oxo-Guo. No significant association was found for CRC diagnosed beyond 5 years of enrollment.

**CONCLUSION:** This novel finding of an inverse and time-dependent relationship between systemic OxS and CRC risk, if further confirmed, may provide a new perspective for revisiting redox-based chemoprevention.

**CONTEXT:** *Background:* Almost all large randomized controlled trials have failed to validate the hypothesis that neutralizing oxidative stress through antioxidant supplementation can lower cancer risk, which has puzzled the public and researchers for decades.

*Key Findings:* A reduced risk for colorectal cancer (CRC) with increasing systemic oxidative stress, measured by two urinary biomarkers of DNA and RNA oxidation, was observed in two large prospective cohort studies in Shanghai, China, and was replicated in an independent cohort in the United States. This association was time-dependent, with the inverse relationship strengthening as the biomarker assessment neared the time of CRC diagnosis.

*Relevance:* Our study, for the first time, suggests an inverse and time-dependent relationship between systemic oxidative stress and CRC development, which, if further confirmed, may provide a new perspective for revisiting redox-based chemoprevention.

## BACKGROUND

It is conventionally believed that antioxidants can lower cancer risk by neutralizing cell-damaging, cancer-causing oxidative stress (OxS). However, despite the compelling biochemical evidence that ties OxS to human carcinogenesis,^1–3^ almost all large randomized controlled trials (RCT) have shown that antioxidant supplementation provides no clear benefit and even increases cancer risk.^4–9^ These seemingly contradictory results have disappointed and puzzled the public and researchers for decades.

It has been suggested that OxS can play dual roles in tumorigenesis— excessive reactive oxygen species (ROS) not only cause *de novo* mutagenesis and tumor progression but also lead to cellular senescence and apoptosis—essentially having both pro-tumor and anti-tumor properties.^10–12^ Emerging evidence from *in vitro* and animal experiments suggests that cancer cells under metabolic stress evolve adaptive mechanisms against OxS through upregulation of cellular antioxidants and antioxidant-scavenging enzymes.^12–14^ By reducing OxS via genetic or pharmacological manipulation in experimental models, cancer cells gain a survival edge, resulting in enhanced growth and metastasis.^15–17^ These experimental advances suggest a shift in the role of OxS from being pro-tumorigenic to anti-tumorigenic over the course of cancer development.^10,17–19^ To our knowledge, this complex effect of OxS at the cellular level in experimental models has not been directly evaluated at the systemic level in a population-based setting.

Unlike previous studies that were often constrained by small sample sizes or retrospective designs, which hindered the prospective evaluation of time-dependent relationships, the present study leverages two large population-based cohorts in Shanghai, China―the Shanghai Women’s Health Study (SWHS) and the Shanghai Men’s Health Study (SMHS)―for the primary analysis,^20,21^ and an independent cohort study in the United States―the Southern Community Cohort Study (SCCS)―for replication.^22^ Systemic OxS was assessed by measuring oxidatively modified guanine nucleosides in urine, specifically 8-oxo-7,8-dihydro-2’-deoxyguanosine (8-oxo-dG) and 7,8-dihydro-8-oxo-guanosine (8-oxo-Guo). Urinary 8-oxo-dG, a well-established biomarker of DNA oxidation, is widely recognized for its reliability in assessing *in vivo* OxS.^23–25^ Meanwhile, 8-oxo-Guo, which reflects RNA oxidation, has gained recognition for its sensitivity in capturing OxS, given RNA’s rapid turnover and greater susceptibility to oxidative damage than DNA.^26–28^ Recent research has highlighted urinary 8-oxo-Guo as a significant biomarker for systemic OxS, complementing 8-oxo-dG by providing additional insights into oxidative damage from various cellular compartments.^28,29^

In this large molecular epidemiological study of colorectal cancer (CRC), we sought to examine whether baseline levels of urinary 8-oxo-dG and 8-oxo-Guo are associated with subsequent risk of CRC and whether the association is time-varying. Additionally, because levels of urinary nucleic acid oxidation markers, particularly 8-oxo-dG, reflect both free radical-induced formation and enzymatic repair of oxidative damage, we further evaluated the potential influence of genetic variations in DNA repair genes on the study results.

## METHODS

### Study Population Parent cohorts

This nested case-control study was conducted in the two Shanghai cohorts, SWHS and SMHS, for the primary analysis, with independent replication of findings in the US cohort, SCCS. Details of the design and methods for SWHS and SMHS have been reported elsewhere.^20,21^ Briefly, 74,941 females aged 40-70 years were recruited to SWHS between 1996 and 2000 (response rate, 92.7%), while 61,480 males aged 40-74 years were recruited to SMHS between 2002 and 2006 (response rate, 74.1%). SCCS is a prospective cohort study conducted across 12 southern US states, enrolling over 85,000 participants aged 40-79 years between 2002 and 2009, with approximately two-thirds being African American (AA).^22^ Baseline exposures were assessed through in-person interviews in the three cohorts, and spot urine samples were collected from 87.7% SWHS, 89.1% SMHS, and 27.6% SCCS participants, after obtaining written informed consent. Protocols for the three cohort studies were approved by institutional review boards for human research at all participating institutes.

### Follow-up and cancer identification

Cohort members in SWHS and SMHS have been followed for occurrence of cancer through a combination of repeated home visits and annual record linkage to Shanghai Tumor Registry and Shanghai Vital Statistics. The diagnosis was verified via reviewing medical records. Incident cancer cases in SCCS were determined through linkage with 12 US state cancer registries and National Death Index mortality records. CRC cases included in this study were cancer-free at baseline and were diagnosed with CRC as the first cancer diagnosis.

### The nested case-control study

During a median follow-up of 15.1 years in SWHS and SMHS, 1938 incident cases of CRC (1035 females and 903 males) with baseline urine samples were identified and included in this study. For each case, one control was randomly selected using incidence-density sampling to individually match on age at baseline (±2 years), sex, date (±30 days) and time (morning/afternoon) of sample collection, and interval since the last meal (±2 hours) among those who provided baseline urine samples and were cancer-free at the time of cancer diagnosis for the index case.

Over 13-year follow-up in SCCS, 285 incident CRC cases were identified among cancer-free participants with baseline urine samples. Each case was matched to two controls using the same criteria, with the inclusion of race. Non-Black or nor-White participants (4.9%) were excluded to avoid low statistical power.

### Laboratory Measurements

Selected biomarkers were measured by liquid chromatography-mass spectrometry (UPLC-MS/MS) at the VUMC Eicosanoid Core Laboratory (**Supplementary Method**).^30,31^ Samples from each case-control pair were analyzed in the same assay batch with a random and blind arrangement. The limit of detection was 0.09 pg/μL for 8-oxo-dG and 0.10 pg/μL for 8-oxo-Guo. Precision of the assay was ±6%, and accuracy was 92.7%. The average within-day and between-day assay coefficients of variation (CV) were lower than 6%, ranging from 0.5% to 16.7%. All measurements were standardized for variations in urinary flow using urine creatinine and expressed as nanograms per milligram of creatinine (ng/mg Cr).

### Statistical Analysis

This study’s primary analyses were performed in SWHS and SMHS, and the independent replication was conducted in SCCS. Conditional logistic regression models, with matched case-control sets as strata, were used to estimate odds ratios (ORs) and 95% confidence intervals (CIs) for CRC risk in association with levels of selected biomarkers. Restricted cubic spline functions with 3 knots were applied to evaluate potential nonlinear effects,^32^ and the exposure and outcome relationship was graphically illustrated. Exposures were categorized based on percentile distributions of biomarker concentrations in controls. ORs for biomarkers at the 10th, 30th, 70th, and 90th percentiles, compared to the 50th percentile (reference), were presented.

Potential confounders adjusted for in multivariable models included age, education, cigarette smoking, alcohol consumption, body mass index (BMI), physical activity, menopausal status (for female participants), regular use of vitamin supplements, regular use of aspirin and other nonsteroidal anti-inflammatory drugs (NSAIDs), Charlson comorbidity score, history of CRC in first-degree relatives, and total energy intake. Stratified analyses by time elapsed from baseline to cancer diagnosis were performed to examine whether the relationship was time varying. Meta-analyses were performed to combine the results of the primary and replication cohorts by using the fixed effects model.

Moreover, to assess potential confounding by genetic variations in DNA repair genes, we selected 35 functional single nucleotide polymorphisms (SNPs) (**Supplementary Method**) and evaluated their associations with urinary 8-oxo-dG and 8-oxo-Guo concentrations and CRC risk in a subset of the study. We further examined whether the association of 8-oxo-dG and 8-oxo-Guo with CRC risk was independent of genetically determined DNA repair capacity. All statistical analyses were conducted using R software (version 4.0.4) with significance defined as two-sided *P* < 0.05.

## RESULTS

### Characteristics of Study Participants

The primary analysis in SWHS and SMHS included 1938 CRC case-control pairs, with the mean age (SD) for cases at diagnosis of 68.3 (9.3) years, and 53.4% were female. Except for a higher likelihood of CRC family history in cases, both groups were comparable in demographics, lifestyle, and known risk factors (**Supplementary Table 1**). This similarity was also observed when analyzing male and female participants separately.

Among 285 CRC cases and 570 matched controls in SCCS, the mean age (SD) for cases at diagnosis was 55.1 (8.8) years, including 67.4% AAs and 33.6% European Americans (EAs). Baseline characteristics were largely comparable between case and control groups (**Supplementary Table 2**).

### Urinary Levels of Nucleic Acid Oxidation Biomarkers Inversely Associated with Subsequent CRC Risk in SWHS and SMHS

Urinary 8-oxo-dG levels were significantly lower in CRC cases compared to their controls **(Supplementary Table 3).** The geometric mean (95% CI) for CRC cases was 8.08 ng/mg Cr (7.87 to 8.29) versus 8.72 ng/mg Cr (8.49 to 8.95) for controls (*P* < 0.001). Urinary 8-oxo-Guo levels were also lower in CRC cases, but not statistically significant. Both urinary 8-oxo-dG and 8-oxo-Guo levels at baseline were significantly and inversely associated with subsequent CRC risk **(Table 1)**. For example, the multivariable-adjusted ORs (95% CI) for CRC at the 10th and 90th percentiles of 8-oxo-dG levels, relative to the median level, were 1.23 (1.07 to 1.41) and 0.82 (0.73 to 0.91), with P for overall association of < 0.001. Similar association patterns were observed when analyzing male and female participants separately **(Supplementary Table 5)**.

**Table 1.**
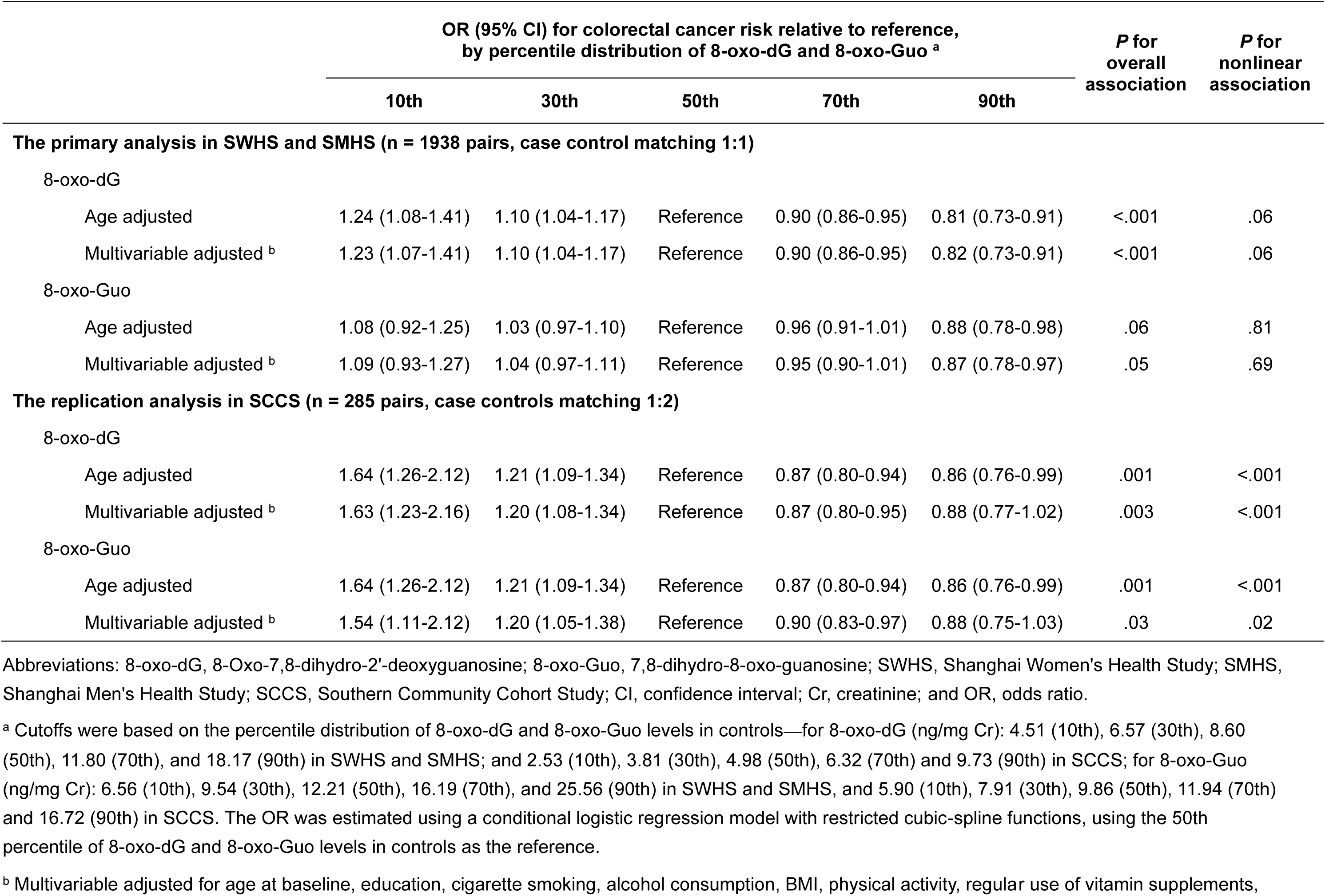
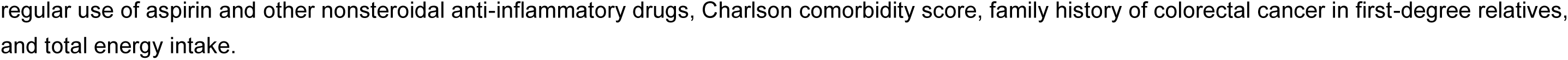
Associations between baseline urinary levels of 8-oxo-dG and 8-oxo-Guo and subsequent risk of colorectal cancer.

### The OxS and CRC Risk Association is Time-Dependent

To evaluate the potential time-varying effect in SWHS and SMHS, CRC cases were categorized based on the timing of cancer diagnosis: within 5 years, between 5 and 9 years, and beyond 9 years after enrollment. Stratified analyses revealed a pronounced temporal pattern in the OxS and CRC risk association, with *P* for heterogeneity of < 0.001 for 8-oxo-dG and 0.07 for 8-oxo-Guo (**Figures 1** and **2**). Baseline urinary 8-oxo-dG and 8-oxo-Guo levels were strongly associated with the risk of CRC diagnosed within 5 years of enrollment (*P* < 0.001 for both biomarkers). ORs (95% CI) for CRC at the 10th and 90th percentiles, relative to the median level of 8-oxo-dG, were 1.87 (1.39 to 2.53) and 0.48 (0.37 to 0.63), respectively, demonstrating an approximate 4-fold difference in risk between the two groups (**Table 2**). Similarly, an approximate 3-fold difference in risk between the two groups was observed for 8-oxo-Guo, with corresponding ORs (95% CI) of 1.46 (1.07 to 1.98) and 0.57 (0.42 to 0.76), respectively. However, no significant association was found for CRC diagnosed beyond 5 years after enrollment.

**Figure 1.**
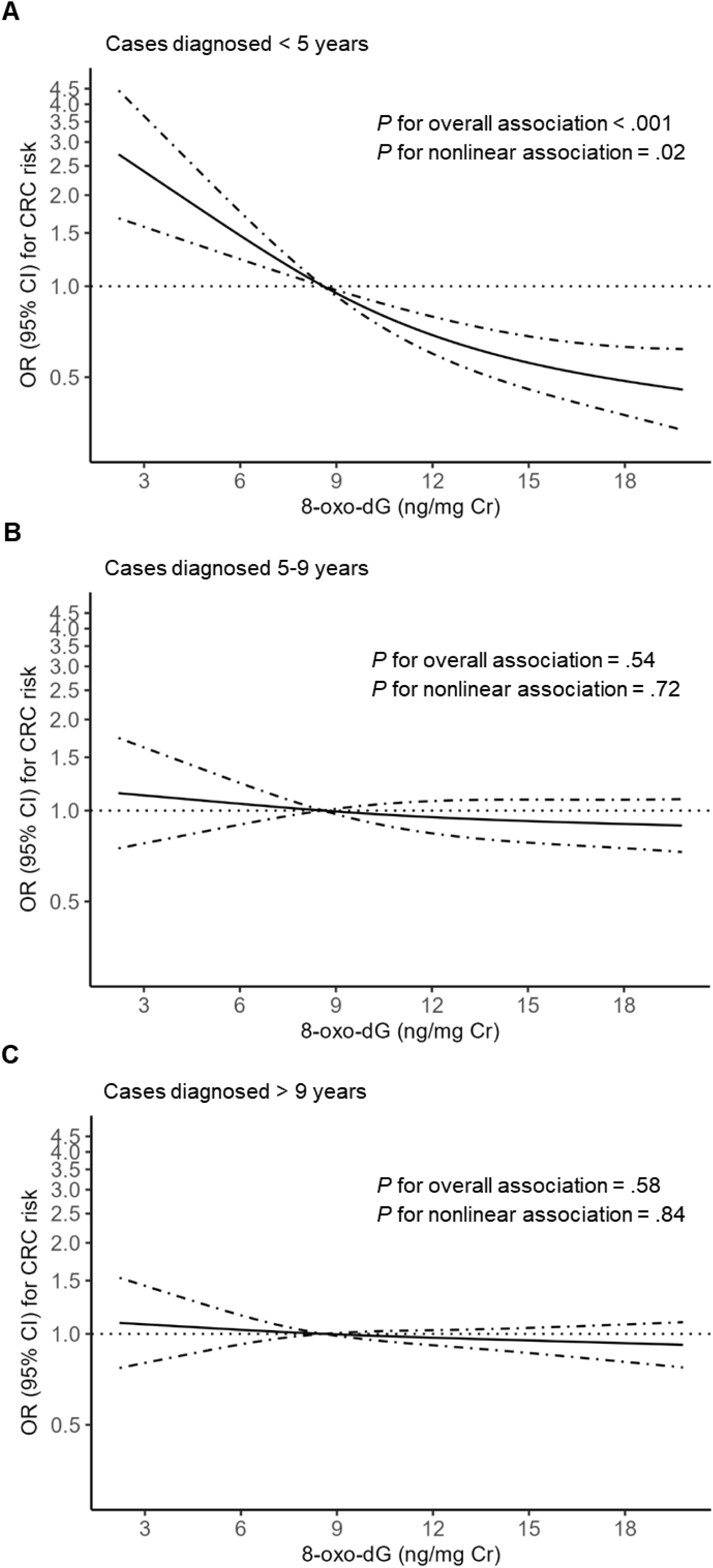
Smoothed plot for multivariable-adjusted ORs for colorectal cancer risk according to urinary levels of 8-oxo-dG, stratified by time interval between enrollment and cancer diagnosis in SWHS and SMHS Colorectal cancer cases were categorized based on the timing of cancer diagnosis: (A) within 5 years, (B) between 5 and 9 years, and (C) beyond 9 years after enrollment. The number of case-control pairs were 429 in group A, 540 in group B, and 969 in group C. ORs were estimated using a conditional logistic regression model with restricted cubic spline functions, using the 50th percentile of 8-oxo-dG levels in controls as the reference. Covariates included in the model are detailed in the footnotes of Table 1. OR values on the *y* axis are shown in log scale. The solid line represents the point estimate, while the dashed lines indicate the 95% CI. *P* for heterogeneity across the three time-interval groups was < 0.001. Abbreviations: 8-oxo-dG denotes 8-Oxo-7,8-dihydro-2’-deoxyguanosine; SWHS, Shanghai Women’s Health Study; SMHS, Shanghai Men’s Health Study; CI, confidence interval; Cr, creatinine; and OR, odds ratio.

**Figure 2.**
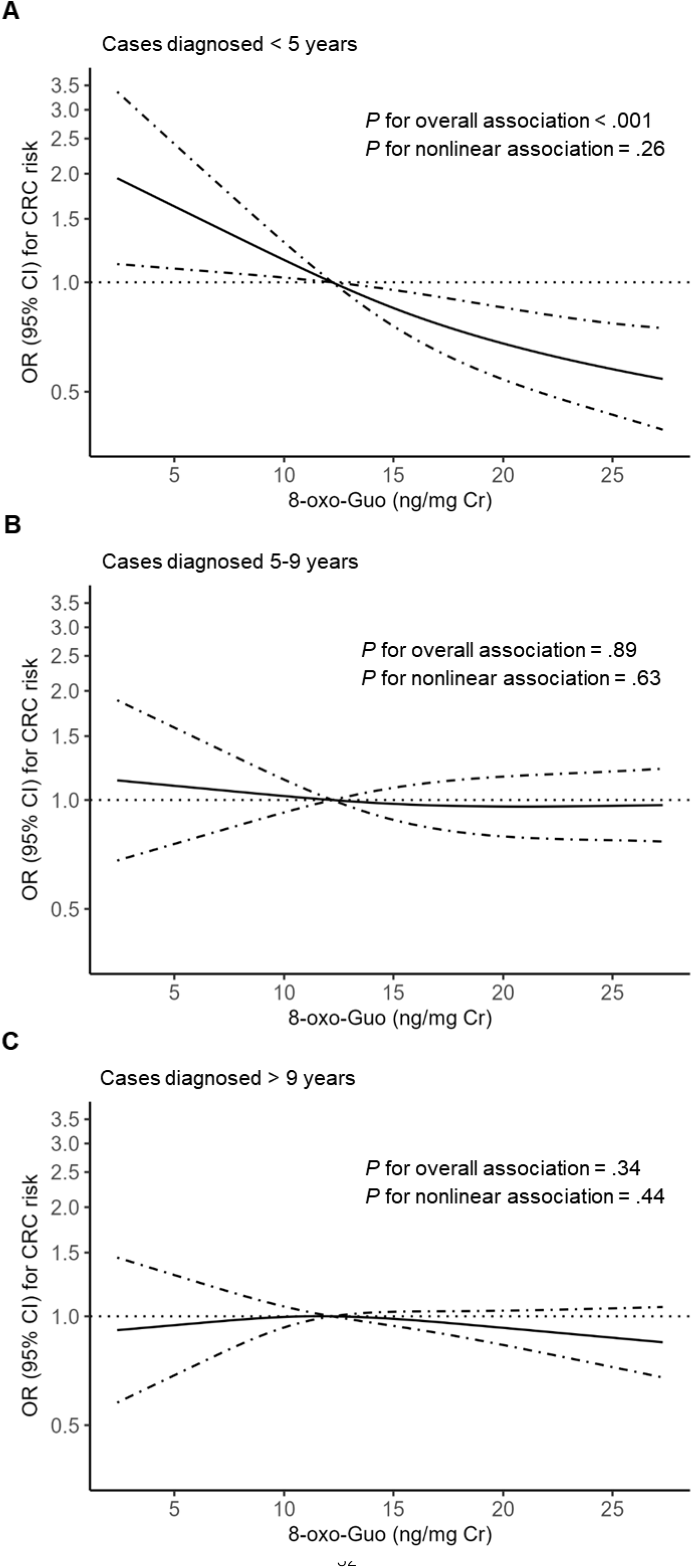
Smoothed plot for multivariable-adjusted ORs for colorectal cancer risk according to urinary levels of 8-oxo-Guo, stratified by time interval between enrollment and cancer diagnosis in SWHS and SMHS Colorectal cancer cases were categorized based on the timing of cancer diagnosis: (A) within 5 years, (B) between 5 and 9 years, and (C) beyond 9 years after enrollment. The number of case-control pairs were 429 in group A, 540 in group B, and 969 in group C. ORs were estimated using a conditional logistic regression model with restricted cubic spline functions, using the 50th percentile of 8-oxo-Guo levels in controls as the reference. Covariates included in the model are detailed in the footnotes of Table 1. OR values on the *y* axis are shown in log scale. The solid line represents the point estimate, while the dashed lines indicate the 95% CI. *P* for heterogeneity across the three time-interval groups was 0.07. Abbreviations: 8-oxo-Guo denotes 7,8-dihydro-8-oxo-guanosine; SWHS, Shanghai Women’s Health Study; SMHS, Shanghai Men’s Health Study; CI, confidence interval; Cr, creatinine; and OR, odds ratio.

**Table 2.**
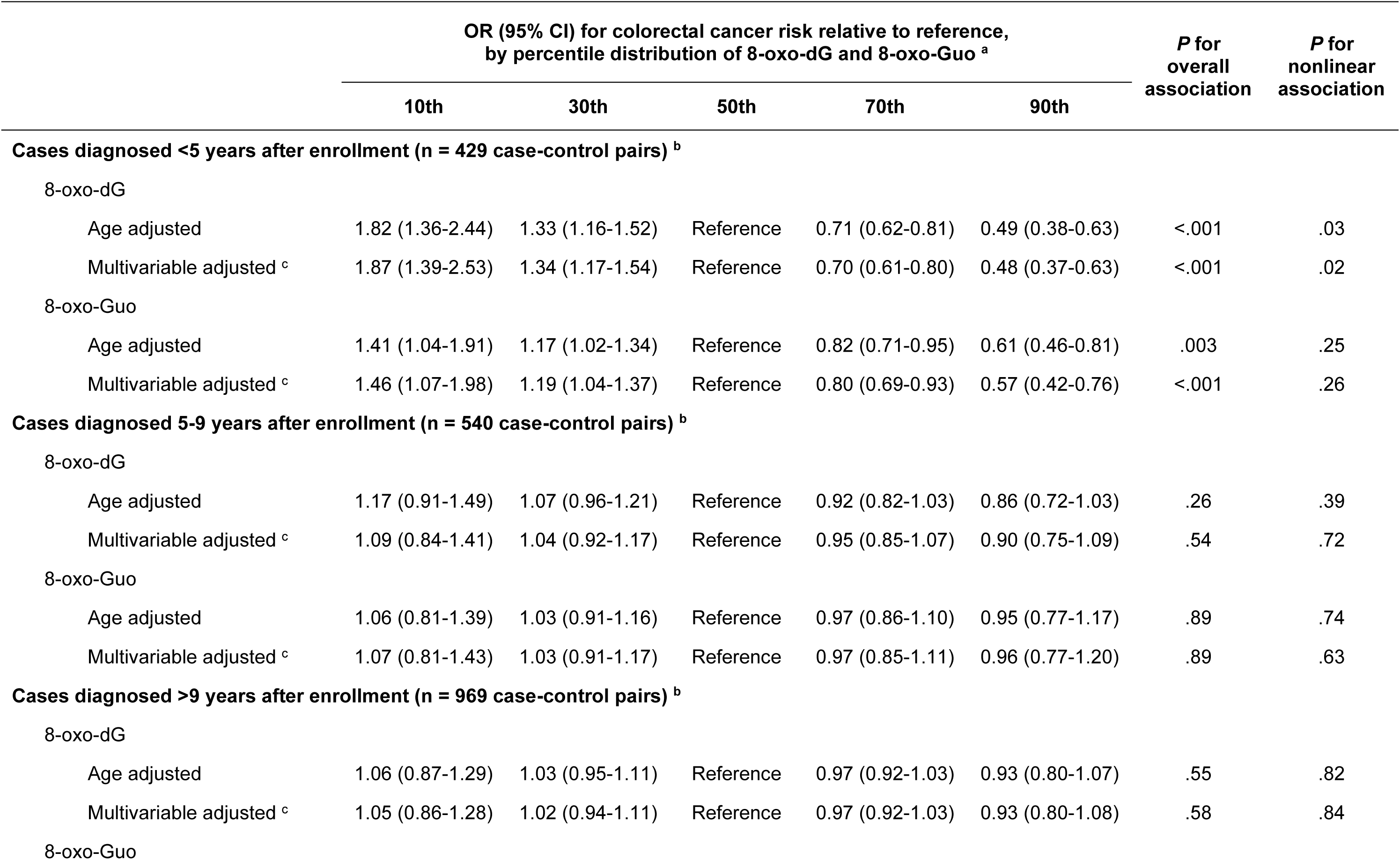
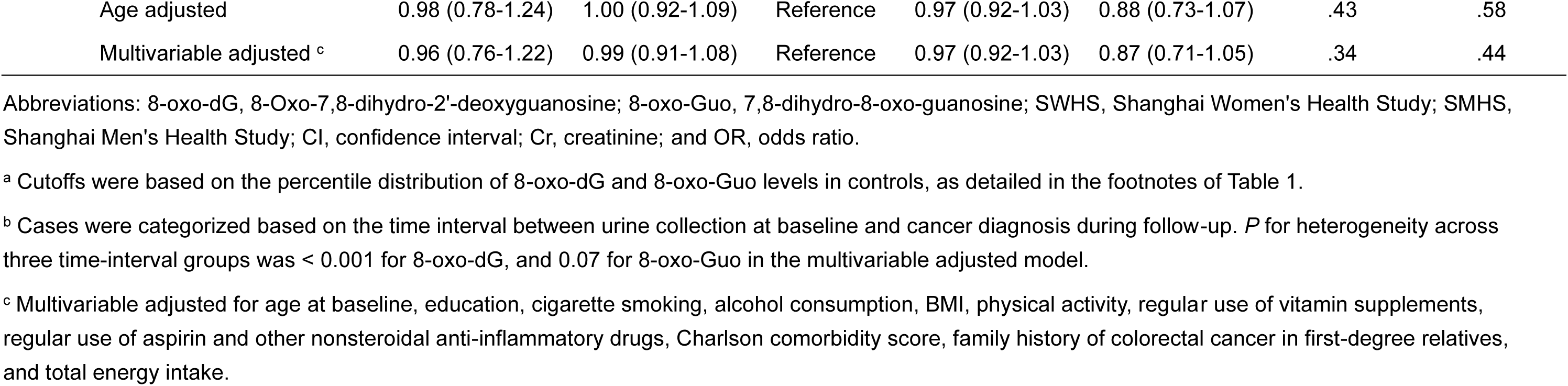
Associations between baseline urinary levels of 8-oxo-dG and 8-oxo-Guo and subsequent risk of colorectal cancer by time interval between urine sample collection at baseline and cancer diagnosis during follow-up in SWHS and SMHS.

The robustness of the study findings was evaluated, revealing that the inverse and time-dependent association remained consistent across various strata by sex, anatomic location and clinical stage of CRC (**Supplementary Tables 7-10**). We also assessed whether the choice of time-interval cutoffs affected the results. The same time-dependent association was observed with finer stratification (data not shown).

### The Association between Nucleic Acid Oxidation Biomarkers and CRC Risk was Independent of Genetically Determined DNA Repair Capacity

As shown in **Supplementary Table 11**, we selected 35 SNPs with potential functional significance from 9 DNA repair genes involved in the formation and metabolism of 8-oxo-dG.^33,34^ Of these, 6 SNPs from 5 genes showed nominal associations with either 8-oxo-dG concentrations (**Supplementary Table 12**) or CRC risk (**Supplementary Table 13**), including one SNP nominally associated with 8-oxo-Guo concentrations. However, after correcting for the false discovery rate, none of these associations remained statistically significant. Further adjustments for these SNPs did not alter the relationship of CRC risk with either 8-oxo-dG or 8-oxo-Guo (**Supplementary Table 14**), suggesting that genetically determined DNA repair capacity is unlikely to significantly skew the study results.

### Replication in SCCS

As demonstrated in SWHS and SMHS, both 8-oxo-dG and 8-oxo-Guo levels in SCCS were also significantly lower in CRC cases than in controls (**Supplementary Table 4**). The smaller number of cases in SCCS (n=285), relative to the Shanghai cohorts (n=1938), limited our ability to conduct stratified analyses by follow-up time, so we only compared the overall effect between the primary and replication cohorts. The inversion association of both OxS biomarkers with CRC risk was replicated in SCCS (**Table 1**). Compared to the median level of biomarkers, the multivariable-adjusted ORs (95% CI) for CRC risk at the 10th and 90th percentiles were 1.63 (1.23 to 2.16) and 0.88 (0.77 to 1.02) for 8-oxo-dG (*P* = 0.003), and 1.54 (1.11 to 2.12) and 0.88 (0.75 to 1.03) for 8-oxo-Guo (*P* = 0.03), respectively. Similar associations were observed when analyzing AA and EA participants separately **(Supplementary Table 6)**.

From meta-analyses pooling the effect estimates from SWHS, SMHS, and SCCS, the corresponding ORs (95% CI) at the 10th and 90th percentiles of biomarker levels were 1.23 (1.22 to 1.24) and 0.82 (0.81 to 0.82) for 8-oxo-dG (*P* < 0.001), and 1.09 (1.08 to 1.10) and 0.87 (0.87 to 0.88) for 8-oxo-Guo (*P* < 0.001), respectively.

## DISCUSSION

This study revealed, for the first time, an inverse and time-varying association between two OxS markers, 8-oxo-dG and 8-oxo-Guo, and CRC risk. Specifically, compared to the control group, CRC patients tended to have lower levels of urinary 8-oxo-dG and 8-oxo-Guo in the later years, but not in the earlier years, of cancer development. This time-varying inverse association remained robust in stratified analyses by sex, anatomical site, and clinical stage of CRC. Furthermore, the inverse association was consistently observed in the independent replication cohort, despite differences in baseline characteristics, including racial ethnicity and risk profiles, between the primary Shanghai cohorts and the replication US cohort.

8-oxo-dG and 8-oxo-Guo have traditionally been a focal point in cancer research, as they represent OxS-induced damage to nucleic acids, a critical process in mutagenesis and carcinogenesis.^12,35,36^ However, emerging evidence suggests a transition in the role of OxS from fostering to suppressing tumor growth across the various phases of cancer evolution.^10,17–19^ Cancer cells are typically more vulnerable to high OxS than normal cells. Increased OxS, if not controlled, can damage macromolecules vital for cell survival, resulting in senescence or apoptosis, thus acting like a tumor suppressor.^37,38^ Cancer cells under metabolic stress undergo metabolic reprogramming.^12–14^ This process activates a variety of antioxidant defense mechanisms to mitigate OxS, thereby promoting further growth and metastasis of cancer.^13,14,19,39^ For example, previous research has demonstrated an increased expression of superoxide dismutase, a key antioxidant enzyme, in CRC patients.^40,41^ The increase was observed both in tumor tissues and in circulation, and it closely correlated with the severity of CRC.^42,43^ Similarly, patients with stage II-IV lung cancer exhibited significantly lower levels of urinary 8-oxo-dG than those with stage I lung cancer,^44^ indicating a decrease in systemic OxS as the malignancy progresses. Our study provides the first population-based evidence on the dynamic relationship between OxS and CRC development over time.

Given that more than two decades can elapse between initiation and clinical emergence of cancer in humans,^45^ individuals diagnosed within the first 5 years of enrollment, as examined in this study, are very likely to have already been in their later phase of cancer development at enrollment. Therefore, the new finding of this study suggests that systemic changes in OxS in the later phase of colorectal tumorigenesis likely occur in parallel with metabolic reprogramming at the original site of cancer. Because we do not fully understand signaling pathways responsible for the systemic change and because this is a new finding, further investigation into the temporal pattern is truly warranted.

Many large RCTs have been conducted to test the hypothesis that lowering OxS through antioxidant supplementation could reduce cancer risk. However, almost all the trials have failed to validate the hypothesis, and some have even shown that antioxidant supplementation increases cancer burden in humans.^4–9^ To our knowledge, the premise of these RCTs primarily stems from survey data suggesting that increasing intake of antioxidants through dietary changes or supplementation is associated with reduced risk of cancer.^46–48^ Supporting evidence from studies directly investigating OxS markers, such as 8-oxo-dG, has been limited and mixed.^49–53^ Both decreased and increased levels of 8-oxo-dG in urine or plasma have been detected in cancer patients in previous studies.^49–51^ The previous efforts had employed a cross-sectional design, comparing cancer-free individuals to cancer cases with biological samples collected at diagnosis and often during cancer treatment.^49–51^ This approach is problematic for investigating alterations related to earlier events. Moreover, most previous studies involved a small sample (<100 cases),^49–51^ and some used biomarkers not validated in both animal and clinical models.^52,53^ More importantly, little attention has been paid to the potential time-dependent effect of OxS.

The present study, to our knowledge, is the largest study of its kind and the first to evaluate the association in a time-dependent manner. The observed inverse and time-dependent relationship between systemic OxS and CRC risk implies that lowering systemic OxS through antioxidant supplementation may not confer benefits and could potentially pose risks for individuals at high risk or with existing precancers. This finding may also provide alternative mechanistic insights into the long-standing question of why antioxidant interventions have led to detrimental outcomes in RCTs among high-risk individuals.^4–6^

Our study has several notable strengths. These include the two-stage approach involving two primary and one replication cohorts, use of well-validated biomarkers, and biomarker quantification by mass spectrometric-based methods, which ensure high scientific rigor regarding measurement accuracy and reproducibility.^31,54^ The large sample size and long-term observation in SWHS and SMHS allowed us to investigate alterations in OxS with CRC progression, although the number of cases in SCCS was insufficient for evaluating this time-varying effect.

Some limitations or concerns should be mentioned. First, urine samples were collected at a single time point. However, it has been demonstrated that levels of 8-oxo-dG and 8-oxo-dG in urine remain stable over time in healthy adults, suggesting that a single measurement can reasonably reflect long-term OxS exposure.^55,56^ Another concern is the possibility of residual confounding, although a wide range of covariates had already been considered in this study. Additionally, it has been suggested that the OxS and cancer risk association may vary by cancer types.^57–59^ Therefore, further investigation into this time-varying effect for various cancer types is clearly warranted.

In conclusion, our study suggests a gradual decline in systemic OxS during the late phases of colorectal tumorigenesis in humans. The time-dependent relationship between systemic OxS and CRC development, if further confirmed, may provide a new prospective for revisiting the redox-based chemoprevention.

## Data Availability

Data described in the article, code book, and analytic codes will be made available upon request pending approval by the data management and sharing committees of SWHS, SMHS and SCCS.

## Author Contributions

Dr. Yang had full access to all the data in the study and take responsibility for the integrity of the data and the accuracy of the data analysis.

*Concept and design:* Yang.

*Acquisition, analysis, or interpretation of data:* All authors.

*Drafting of the manuscript:* Zhao, Nogueira, Chen, Yang.

*Critical revision of the manuscript for important intellectual content:* All authors.

*Statistical analysis:* Zhao, Chen, Yang.

*Obtained funding:* Shu, Zheng, Chen, Yang.

*Administrative, technical, or material support:* Zhao, Nogueira, Milne, Cai, Chen, Yang.

*Supervision:* Yang.

## Conflict of Interest Disclosures

None reported.

## Funding/Support

This work was supported by the National Institutes of Health (grant numbers R01CA237895, R01CA122364, UM1CA182910, UM1CA173640 and U01CA202979) and was in part supported by the Vanderbilt-Ingram Cancer Center (P30CA068485).

## Role of the Funder/Sponsor

The funders had no role in the design and conduct of the study; collection, management, analysis, or interpretation of the data; preparation, review, or approval of the manuscript; or the decision to submit the manuscript for publication.

## Additional Contributions

We are grateful to the participants and research staff of the Shanghai Women’s Health Study, the Shanghai Men’s Health Study, and the Southern Community Cohort Study for their contributions to the study. We thank Jie Wu, Regina Courtney and Rodica Cal-Chris for contributions to sample preparation at the Survey and Biospecimen Shared Resources. Further, we thank Professor Henrik Enghusen Poulsen of the Laboratory of Clinical Pharmacology at the University of Copenhagen for his graciousness in sharing the laboratory analysis protocol and breadth of knowledge on the subject with us. These individuals were not compensated.

## Notes

### Competing Interest Statement

The authors have declared no competing interest.

### Author Declarations

Vanderbilt University Medical Center IRB #191648 "Time-dependent and bidirectional effect of oxidative stress: a missing piece of the free radical theory of cancer and its potential implications" The designee determined the study does not qualify as "human subject" research per 46.102(f)(2).

