## Supplemental data for "Time-dependent relationship between urinary biomarkers of nucleic acid oxidation and colorectal cancer risk"

### Supplementary Online Content


1. **Supplementary Method**
2. **Supplementary Table 1.** Baseline characteristics of colorectal cancer cases and matched controls in SWHS and SMHS
3. **Supplementary Table 2.** Baseline characteristics of colorectal cancer cases and matched controls in SCCS
4. **Supplementary Table 3.** Urinary levels of 8-oxo-dG and 8-oxo-Guo in colorectal cancer cases and controls from SWHS and SMHS
5. **Supplementary Table 4.** Urinary levels of 8-oxo-dG and 8-oxo-Guo in colorectal cancer cases and controls from SCCS
6. **Supplementary Table 5.** Associations between baseline urinary levels of 8-oxo-dG and 8-oxo-Guo and subsequent risk of colorectal cancer by sex (cohort) in SWHS and SMHS
7. **Supplementary Table 6.** Associations between baseline urinary levels of 8-oxo-dG and 8-oxo-Guo and subsequent risk of colorectal cancer by race in SCCS
8. **Supplementary Table 7.** Associations between baseline urinary levels of 8-oxo-dG and 8-oxo-Guo and subsequent risk of colorectal cancer by time interval between urine sample collection at baseline and cancer diagnosis during follow-up in SWHS
9. **Supplementary Table 8.** Associations between baseline urinary levels of 8-

oxo-dG and 8-oxo-Guo and subsequent risk of colorectal cancer by time interval between urine sample collection at baseline and cancer diagnosis during follow-up in SMHS

10. **Supplementary Table 9.** Associations between baseline urinary levels of 8-oxo-dG and 8-oxo-Guo and subsequent risk of colorectal cancer in SWHS and SMHS, stratified by cancer anatomical sites and by time interval between urine sample collection at baseline and cancer diagnosis during follow-up
11. **Supplementary Table 10.** Associations between baseline urinary levels of 8-oxo-dG and 8-oxo-Guo and subsequent risk of colorectal cancer in SWHS and SMHS, stratified by cancer stages and by time interval between urine sample collection at baseline and cancer diagnosis during follow-up
12. **Supplementary Table 11.** Selected functional SNPs in DNA repair genes
13. **Supplementary Table 12.** Associations of selected functional SNPs in DNA repair genes with urinary levels of 8-oxo-dG and 8-oxo-Guo among controls from SWHS and SMHS
14. **Supplementary Table 13.** Associations between selected functional SNPs in DNA repair genes and colorectal cancer risk in SWHS and SMHS
15. **Supplementary Table 14.** Associations between baseline urinary levels of 8-oxo-dG and 8-oxo-Guo and subsequent risk of colorectal cancer overall and by time interval between urine sample collection at baseline and cancer diagnosis during follow-up in SWHS and SMHS, after further adjustment for selected functional SNPs in DNA repair genes

### 16. References

### Supplementary Method

#### Laboratory Measurements

Concentrations of 8-oxo-dG and 8-oxo-Guo in urine were measured at the VUMC Eicosanoid Core Laboratory.[1-3] 8-oxo-dG, $^{13}C,^{15}N_2$-8-oxo-dG and 8-oxo-Guo were purchased from Santa Cruz Biotechnology (Dallas, TX USA). All solvents were of LC/MS grade and were purchased from ThermoFisher Scientific (Waltham, MA USA).

Frozen urine samples were thawed and a 50µL aliquot was removed. The sample was diluted with DI water (190µL) and the internal standard solution (2ng in 10µL in ethanol). The samples were mixed by vortexing and then passed through a Costar® Spin-X filter (cellulose acetate membrane, pore size 0.45µm, Corning Inc., Corning, NY USA) while centrifuging at 13,000G for 3min.

8-oxo-dG and 8-oxo-Guo was then quantified using liquid chromatography-mass spectrometry (UPLC-MS/MS). UPLC-MS/MS was performed on a Waters Xevo TQ-XS triple quadrupole mass spectrometer coupled with an Acquity I-class UPLC system (Waters Corporation, Milford, MA, USA). The capillary voltage was set to 2.5kV, the cone voltage was 15kV, and the collision energy was 10. Analytes were separated using a Waters Acquity UPLC BEH C18 column (2.1 x 50mm, 1.7µm) and a gradient mobile phase system where Mobile Phase A (MPA) was water containing 0.1% acetic acid and Mobile Phase B (MPB) was methanol. The flow rate was 0.250mL/min. The gradient progressed from 99% MPA to 50% MPA over 7 minutes; the column was washed using 5% MPA for 3 minutes and then returned to initial conditions. The MS was operated using multiple reaction monitoring in the negative ion mode and the transitions ions for 8-oxo-dG, $^{13}C,^{15}N_2$-8-oxo-dG and 8-oxo-Guo were 284 → 168, 287 → 171 and 300 → 168 respectively. Analyte quantification was carried out based on the ratio of the analyte to internal standard peak height. Instrument control and data acquisition utilized MassLynx V4.2; integration and quantitation used TargetLynx 4.2.

Urinary creatinine levels were measured using a test kit from Enzo Life Sciences (Farmingdale, NY USA) according to manufacturer instructions. All urinary measurements for 8-oxo-dG and 8-oxo-Guo were calculated in ng/mL and were then standardized for variations in urinary flow using urine creatinine and expressed as nanograms per milligram of creatinine (ng/mg Cr).

Samples from each case-control pair were analyzed in the same assay batch with a random and blind arrangement. The limit of detection was 0.09 pg/μL for 8-oxo-dG and 0.10 pg/μL for 8-oxo-Guo. Precision of the assay was ± 6%, and accuracy was 92.7%. The average within-day and between-day assay coefficients of variation (CV) were lower than 6%, ranging from 0.5% to 16.7%.

**DNA Repair Genes and Polymorphisms Selection**

Genetic variants data in this study were available for 3256 participants in SWHS and SMHS, including 1711 cases and 1545 controls, generated by previous studies via Affymetrix SNP array 6.0 (Thermo Fisher Scientific, Waltham, Massachusetts), Illumina 550k Array Omni Express, or Illumina Omni Express (Illumina, Inc., San

Diego, California).[4,5] Detailed information on methodologies for DNA extraction, single nucleotide polymorphism (SNP) genotyping, and imputation have been described elsewhere.[6] All genetic variants data passed stringent quality control and were imputed using 1000 Genomes Project data as a reference panel.

We selected 9 DNA repair genes that have been characterized to be involved in the metabolism of 8-oxo-dG from formation to urinary excretion.[7,8] After excluding SNPs with minor allele frequency < 0.005 and imputation quality metric < 0.3, a total of 35 SNPs with potentially functional significance were selected in these genes, most of which were in the promoter regions, exons, or splice-donor or splice-acceptor sites in introns, based on the literature.[9-29] The selection criteria were that these SNPs were demonstrated to have proven biological activity, causing changes in the function or expression of the encoded protein. The relevant data on genes and selected SNPs are shown in **Supplementary Table 11**.

**Supplementary Table 1. Baseline characteristics of colorectal cancer cases and matched controls in SWHS and SMHS**

| Characteristics | All | | | Females in SWHS | | | Males in SMHS | | |
|---|---|---|---|---|---|---|---|---|---|
| | Cases (n = 1938) | Controls (n = 1938) | *P* value [a] | Cases (n = 1035) | Controls (n = 1035) | *P* value [a] | Cases (n = 903) | Controls (n = 903) | *P* value [a] |
| Age (year), mean (SD) | 59.4 (9.2) | 59.2 (9.2) | .74 | 57.5 (8.8) | 57.5 (8.9) | .98 | 61.5 (9.2) | 61.3 (9.2) | .63 |
| Education, high school and above, n (%) | 865 (44.6) | 844 (43.6) | .52 | 361 (34.9) | 342 (33.0) | .40 | 504 (55.8) | 502 (55.6) | .96 |
| Alcohol consumption, n (%) | 334 (17.2) | 346 (17.9) | .64 | 22 (2.1) | 26 (2.5) | .66 | 312 (34.6) | 320 (35.4) | .73 |
| Cigarette smoking, n (%) | 605 (31.2) | 628 (32.4) | .45 | 29 (2.8) | 38 (3.7) | .32 | 576 (63.8) | 590 (65.3) | .52 |
| Family history of colorectal cancer, n (%) | 65 (3.4) | 36 (1.9) | .005 | 38 (3.7) | 19 (1.8) | .02 | 27 (3.0) | 17 (1.9) | .17 |
| Postmenopausal, n (%) | NA | NA | NA | 749 (72.4) | 759 (73.3) | .68 | NA | NA | NA |
| Physical activity (MET-h/wk), mean (SD) | 89.0 (44.0) | 88.8 (47.3) | .27 | 108.2 (41.5) | 109.7 (45.8) | .91 | 67.1 (35.7) | 64.7 (36.3) | .09 |
| Body mass index ($kg/m^2$), mean (SD) | 24.4 (3.3) | 24.2 (3.4) | .06 | 24.5 (3.4) | 24.6 (3.6) | .54 | 24.3 (3.2) | 23.7 (3.0) | <.001 |
| Use of vitamin supplements, n (%) | 348 (18.0) | 318 (16.4) | .22 | 189 (18.3) | 185 (17.9) | .86 | 159 (17.6) | 133 (14.7) | .11 |
| Use of aspirin and other NSAIDs, n (%) | 134 (6.9) | 115 (5.9) | .24 | 20 (1.9) | 31 (3.0) | .16 | 114 (12.6) | 84 (9.3) | .03 |
| Total energy intake (kcal/d), mean (SD) | 1775.6 (439.8) | 1786.4 (475.0) | .95 | 1673.5 (394.1) | 1693.9 (428.1) | .45 | 1892.6 (460.1) | 1892.5 (503.4) | .55 |
| Charlson comorbidity index, mean (SD) | 0.4 (0.9) | 0.4 (0.8) | .96 | 0.3 (0.7) | 0.3 (0.8) | .06 | 0.6 (1.0) | 0.5 (0.9) | .06 |

Abbreviations: SWHS, Shanghai Women's Health Study; SMHS, Shanghai Men's Health Study; case, study participant who had developed colorectal cancer during follow-up; control, study participant who remained cancer free; MET, metabolic equivalent; NSAID, nonsteroidal anti-inflammatory drug; NA, not applicable.

[a] Wilcoxon rank sum test was used for continuous variables and chi-square test for categorical variables.

**Supplementary Table 2. Baseline characteristics of colorectal cancer cases and matched controls in SCCS**

| Characteristics | All | | | African | | | European | | |
|---|---|---|---|---|---|---|---|---|---|
| | Cases (n = 285) | Controls (n = 570) | *P* value [a] | Cases (n = 192) | Controls (n = 384) | *P* value [a] | Cases (n = 93) | Controls (n = 186) | *P* value [a] |
| Age (year), mean (SD) | 55.1 (8.8) | 55.0 (8.7) | .84 | 54.6 (8.8) | 54.4 (8.7) | .79 | 56.1 (8.7) | 56.1 (8.6) | .98 |
| Education, high school and above, n (%) | 181 (63.5) | 359 (63.0) | .89 | 115 (59.9) | 231 (60.2) | .99 | 66 (71.0) | 128 (68.8) | .82 |
| Alcohol consumption, n (%) | 237 (83.2) | 478 (83.9) | .99 | 165 (85.9) | 325 (84.6) | .82 | 72 (77.4) | 153 (82.3) | .64 |
| Cigarette smoking, n (%) | 180 (63.2) | 371 (65.1) | .73 | 116 (60.4) | 243 (63.3) | .62 | 64 (68.8) | 128 (68.8) | .99 |
| Family history of colorectal cancer, n (%) | 34 (11.9) | 43 (7.5) | .04 | 18 (9.4) | 26 (6.8) | .34 | 16 (17.2) | 17 (9.1) | .07 |
| Gender, male (%) | 112 (39.3) | 224 (39.3) | .99 | 80 (41.7) | 160 (41.7) | .99 | 32 (34.4) | 64 (34.4) | .99 |
| Physical activity (MET-h/wk), mean (SD) | 160.0 (136.5) | 160.7 (140.5) | .90 | 156.5 (142.4) | 152.9 (141.8) | .71 | 167.4 (123.6) | 176.8 (136.9) | .70 |
| Body mass index (kg/m$^2$), mean (SD) | 30.7 (7.9) | 31.2 (7.6) | .23 | 30.1 (7.4) | 31.2 (8.0) | .14 | 31.8 (8.7) | 31.1 (6.9) | .87 |
| Use of vitamin supplements, n (%) | 116 (40.7) | 249 (43.7) | .49 | 78 (40.6) | 156 (40.6) | .99 | 38 (40.9) | 93 (50.0) | .25 |
| Use of aspirin and other NSAIDs, n (%) | 32 (11.2) | 73 (12.8) | .59 | 21 (10.9) | 50 (13.0) | .56 | 11 (11.8) | 23 (12.4) | .99 |
| Total energy intake (kcal/d), mean (SD) | 2596.9 (1540.5) | 2547.2 (1439.6) | .89 | 2726.8 (1591.5) | 2692.5 (1524.6) | .87 | 2334.3 (1403.7) | 2262.3 (1210.5) | .86 |
| Charlson comorbidity index, mean (SD) | 2.0 (1.5) | 2.1 (1.4) | .30 | 1.9 (1.5) | 2.1 (1.4) | .10 | 2.3 (1.6) | 2.2 (1.4) | .54 |

Abbreviations: SCCS, Southern Community Cohort Study; case, study participant who had developed colorectal cancer; control, study participant who remained cancer free; MET, metabolic equivalent; NSAID, nonsteroidal anti-inflammatory drug.

[a] Wilcoxon rank sum test was used for continuous variables and chi-square test for categorical variables

**Supplementary Table 3. Urinary levels of 8-oxo-dG and 8-oxo-Guo in colorectal cancer cases and controls from SWHS and SMHS**

| | Case (ng/mg Cr) | | Control (ng/mg Cr) | | Difference b/w case and control [a] | *P* value [b] |
|---|---|---|---|---|---|---|
| | Geometric mean | 95% CI | Geometric mean | 95% CI | | |
| **All (n = 1938 pairs, case control matching 1:1)** | | | | | | |
| 8-oxo-dG | 8.08 | (7.87-8.29) | 8.72 | (8.49-8.95) | -7.3% | <.001 |
| 8-oxo-Guo | 11.79 | (11.44-12.15) | 12.13 | (11.77-12.50) | -2.8% | .18 |
| **Females (n = 1035 pairs, case control matching 1:1)** | | | | | | |
| 8-oxo-dG | 6.78 | (6.56-7.01) | 7.37 | (7.13-7.62) | -8.0% | <.001 |
| 8-oxo-Guo | 9.59 | (9.25-9.95) | 9.87 | (9.51-10.24) | -2.8% | .30 |
| **Males (n = 903 pairs, case control matching 1:1)** | | | | | | |
| 8-oxo-dG | 9.86 | (9.49-10.25) | 10.56 | (10.16-10.98) | -6.6% | .005 |
| 8-oxo-Guo | 14.93 | (14.29-15.60) | 15.36 | (14.70-16.05) | -2.8% | .38 |

Abbreviations: 8-oxo-dG, 8-Oxo-7,8-dihydro-2'-deoxyguanosine; 8-oxo-Guo, 7,8-dihydro-8-oxo-guanosine; SWHS, Shanghai Women's Health Study; SMHS, Shanghai Men's Health Study; case, study participant who had developed colorectal cancer during follow-up; control, study participant who remained cancer free.

[a] Difference (%) = (geometric mean of biomarker in case − geometric mean of biomarker in control)/geometric mean of biomarker in control.

[b] Derived from a paired *t* test.

**Supplementary Table 4. Urinary levels of 8-oxo-dG and 8-oxo-Guo in colorectal cancer cases and controls from SCCS**

| | Case (ng/mg Cr) | | Control (ng/mg Cr) | | Difference b/w case and control [a] | *P* value [b] |
|---|---|---|---|---|---|---|
| | Geometric mean | 95% CI | Geometric mean | 95% CI | | |
| **All (n = 285 pairs, case controls matching 1:2)** | | | | | | |
| 8-oxo-dG | 4.52 | (4.21-4.86) | 5.19 | (4.93-5.45) | -12.9% | .003 |
| 8-oxo-Guo | 9.36 | (8.86-9.89) | 10.02 | (9.64-10.42) | -6.6% | .03 |
| **African Americans (n = 192 pairs, case controls matching 1:2)** | | | | | | |
| 8-oxo-dG | 4.21 | (3.86-4.59) | 4.91 | (4.62-5.22) | -14.3% | .008 |
| 8-oxo-Guo | 8.65 | (8.07-9.27) | 9.63 | (9.17-10.11) | -10.2% | .01 |
| **European Americans (n = 93 pairs, case controls matching 1:2)** | | | | | | |
| 8-oxo-dG | 5.25 | (4.65-5.93) | 5.83 | (5.34-6.36) | -9.9% | .19 |
| 8-oxo-Guo | 11.01 | (10.10-12.00) | 10.90 | (10.25-11.59) | 1.0% | .88 |

Abbreviations: 8-oxo-dG, 8-Oxo-7,8-dihydro-2'-deoxyguanosine; 8-oxo-Guo, 7,8-dihydro-8-oxo-guanosine; SCCS, Southern Community Cohort Study; case, study participant who had developed colorectal cancer during follow-up; control, study participant who remained cancer free.

[a] Difference (%) = (geometric mean of biomarker in case − geometric mean of biomarker in control)/geometric mean of biomarker in control.

[b] Derived from a paired *t* test.

**Supplementary Table 5. Associations between baseline urinary levels of 8-oxo-dG and 8-oxo-Guo and subsequent risk of colorectal cancer by sex (cohort) in SWHS and SMHS**

| | OR (95% CI) for colorectal cancer risk relative to reference, by percentile distribution of 8-oxo-dG and 8-oxo-Guo [a] | | | | | *P* for overall association | *P* for nonlinear association |
|---|---|---|---|---|---|---|---|
| | 10th | 30th | 50th | 70th | 90th | | |
| **Female participants (n = 1035 pairs, case control matching 1:1)** | | | | | | | |
| 8-oxo-dG | | | | | | | |
| Age adjusted | 1.27 (1.06-1.54) | 1.11 (1.03-1.19) | Reference | 0.92 (0.86-0.97) | 0.82 (0.67-1.01) | .004 | .14 |
| Multivariable adjusted [b] | 1.25 (1.03-1.51) | 1.10 (1.02-1.18) | Reference | 0.92 (0.86-0.98) | 0.82 (0.66-1.01) | .009 | .21 |
| 8-oxo-Guo | | | | | | | |
| Age adjusted | 1.12 (0.91-1.38) | 1.05 (0.98-1.12) | Reference | 0.95 (0.90-1.00) | 0.87 (0.71-1.05) | .19 | .65 |
| Multivariable adjusted [b] | 1.11 (0.90-1.37) | 1.04 (0.97-1.12) | Reference | 0.96 (0.91-1.01) | 0.87 (0.72-1.06) | .25 | .69 |
| **Male participants (n = 903 pairs, case control matching 1:1)** | | | | | | | |
| 8-oxo-dG | | | | | | | |
| Age adjusted | 1.19 (1.00-1.42) | 1.09 (1.00-1.18) | Reference | 0.90 (0.82-0.99) | 0.80 (0.69-0.94) | .02 | .24 |
| Multivariable adjusted [b] | 1.16 (0.96-1.39) | 1.07 (0.98-1.17) | Reference | 0.91 (0.83-1.00) | 0.82 (0.70-0.96) | .05 | .38 |
| 8-oxo-Guo | | | | | | | |
| Age adjusted | 1.03 (0.85-1.25) | 1.01 (0.93-1.11) | Reference | 0.98 (0.88-1.08) | 0.90 (0.75-1.08) | .25 | .81 |
| Multivariable adjusted [b] | 1.04 (0.85-1.26) | 1.02 (0.93-1.11) | Reference | 0.97 (0.88-1.08) | 0.89 (0.74-1.07) | .22 | .84 |

Abbreviations: 8-oxo-dG, 8-Oxo-7,8-dihydro-2'-deoxyguanosine; 8-oxo-Guo, 7,8-dihydro-8-oxo-guanosine; SWHS, Shanghai Women's Health Study; SMHS, Shanghai Men's Health Study; CI, confidence interval; Cr, creatinine; OR, odds ratio.

[a] Cutoffs were based on the percentile distribution of 8-oxo-dG and 8-oxo-Guo levels in controls—for 8-oxo-dG (ng/mg Cr): 4.51 (10th), 6.57 (30th), 8.60 (50th), 11.80 (70th), and 18.17 (90th); and for 8-oxo-Guo (ng/mg Cr): 6.56 (10th), 9.54 (30th), 12.21 (50th), 16.19 (70th), and 25.56 (90th). The OR was estimated by using a conditional logistic regression model with restricted cubic-spline functions, with the 50th percentile of 8-oxo-dG and 8-oxo-Guo levels in controls treated as the reference.

[b] Multivariable adjusted for age at baseline, education, cigarette smoking, alcohol consumption, BMI, physical activity, menopausal status (for female participants),

regular use of vitamin supplements, regular use of aspirin and other nonsteroidal anti-inflammatory drugs, Charlson comorbidity score, family history of colorectal cancer in first-degree relatives, and total energy intake.

**Supplementary Table 6. Associations between baseline urinary levels of 8-oxo-dG and 8-oxo-Guo and subsequent risk of colorectal cancer by race in SCCS**

| | OR (95% CI) for colorectal cancer risk relative to reference, by percentile distribution of 8-oxo-dG and 8-oxo-Guo [a] | | | | | *P* for overall association | *P* for nonlinear association |
|---|---|---|---|---|---|---|---|
| | 10th | 30th | 50th | 70th | 90th | | |
| **African Americans (n = 192 pairs, case controls matching 1:2)** | | | | | | | |
| 8-oxo-dG | | | | | | | |
| Age adjusted | 1.59 (1.17-2.16) | 1.22 (1.07-1.40) | Reference | 0.87 (0.80-0.95) | 0.83 (0.71-0.98) | .01 | .007 |
| Multivariable adjusted [b] | 1.54 (1.11-2.14) | 1.21 (1.05-1.39) | Reference | 0.88 (0.80-0.97) | 0.85 (0.72-1.01) | .03 | .02 |
| 8-oxo-Guo | | | | | | | |
| Age adjusted | 1.59 (1.17-2.16) | 1.22 (1.07-1.40) | Reference | 0.87 (0.80-0.95) | 0.83 (0.71-0.98) | .01 | .007 |
| Multivariable adjusted [b] | 1.46 (1.01-2.12) | 1.20 (1.01-1.43) | Reference | 0.89 (0.80-0.98) | 0.81 (0.66-1.00) | .06 | .12 |
| **European Americans (n = 93 pairs, case controls matching 1:2)** | | | | | | | |
| 8-oxo-dG | | | | | | | |
| Age adjusted | 1.53 (1.02-2.30) | 1.18 (1.01-1.37) | Reference | 0.89 (0.79-1.01) | 0.96 (0.72-1.26) | .10 | .03 |
| Multivariable adjusted [b] | 1.54 (0.96-2.48) | 1.18 (0.98-1.41) | Reference | 0.90 (0.77-1.04) | 0.99 (0.73-1.36) | .14 | .05 |
| 8-oxo-Guo | | | | | | | |
| Age adjusted | 1.53 (1.02-2.30) | 1.18 (1.01-1.37) | Reference | 0.89 (0.79-1.01) | 0.96 (0.72-1.26) | .10 | .03 |
| Multivariable adjusted [b] | 1.53 (0.84-2.79) | 1.15 (0.94-1.42) | Reference | 0.94 (0.84-1.06) | 1.05 (0.80-1.39) | .29 | .12 |

Abbreviations: 8-oxo-dG, 8-Oxo-7,8-dihydro-2'-deoxyguanosine; 8-oxo-Guo, 7,8-dihydro-8-oxo-guanosine; SCCS, Southern Community Cohort Study; CI, confidence interval; Cr, creatinine; OR, odds ratio.

[a] Cutoffs were based on the percentile distribution of 8-oxo-dG and 8-oxo-Guo levels in controls—for 8-oxo-dG: 2.53 (10th), 3.81 (30th), 4.98 (50th), 6.32 (70th) and 9.73 (90th); and for 8-oxo-Guo: 5.90 (10th), 7.91 (30th), 9.86 (50th), 11.94 (70th) and 16.72 (90th). The OR was estimated by using a conditional logistic regression model with restricted cubic-spline functions, with the 50th percentile of 8-oxo-dG and 8-oxo-Guo levels in controls treated as the reference.

[b] Multivariable adjusted for age at baseline, education, cigarette smoking, alcohol consumption, BMI, physical activity, regular use of vitamin supplements, regular use of aspirin and other nonsteroidal anti-inflammatory drugs, Charlson comorbidity score, family history of colorectal cancer in first-degree relatives, and total energy intake.

**Supplementary Table 7. Associations between baseline urinary levels of 8-oxo-dG and 8-oxo-Guo and subsequent risk of colorectal cancer by time interval between urine sample collection at baseline and cancer diagnosis during follow-up in SWHS [a]**

| | OR (95% CI) for colorectal cancer risk relative to reference, by percentile distribution of 8-oxo-dG and 8-oxo-Guo [b] | | | | | *P* for overall association | *P* for nonlinear association |
|---|---|---|---|---|---|---|---|
| | 10th | 30th | 50th | 70th | 90th | | |
| **Cases diagnosed <5 years after enrollment (n = 185 pairs, case control matching 1:1)** | | | | | | | |
| 8-oxo-dG | | | | | | | |
| Age adjusted | 1.45 (0.93-2.26) | 1.22 (1.04-1.45) | Reference | 0.68 (0.54-0.86) | 0.30 (0.12-0.71) | .003 | .65 |
| Multivariable adjusted [c] | 1.37 (0.85-2.22) | 1.21 (1.01-1.45) | Reference | 0.66 (0.51-0.85) | 0.26 (0.10-0.66) | .005 | .43 |
| 8-oxo-Guo | | | | | | | |
| Age adjusted | 1.07 (0.67-1.70) | 1.07 (0.90-1.26) | Reference | 0.81 (0.66-0.99) | 0.42 (0.18-0.98) | .12 | .33 |
| Multivariable adjusted [c] | 1.11 (0.68-1.82) | 1.08 (0.90-1.29) | Reference | 0.82 (0.65-1.02) | 0.45 (0.17-1.13) | .20 | .48 |
| **Cases diagnosed 5-9 years after enrollment (n = 216 pairs, case control matching 1:1)** | | | | | | | |
| 8-oxo-dG | | | | | | | |
| Age adjusted | 1.22 (0.78-1.89) | 1.09 (0.92-1.28) | Reference | 0.92 (0.72-1.18) | 0.80 (0.32-2.00) | .55 | .76 |
| Multivariable adjusted [c] | 1.22 (0.75-1.98) | 1.09 (0.91-1.31) | Reference | 0.91 (0.70-1.17) | 0.77 (0.30-1.98) | .56 | .82 |
| 8-oxo-Guo | | | | | | | |
| Age adjusted | 1.13 (0.68-1.87) | 1.03 (0.85-1.24) | Reference | 1.06 (0.91-1.23) | 1.39 (0.84-2.32) | .36 | .32 |
| Multivariable adjusted [c] | 1.16 (0.66-2.03) | 1.04 (0.84-1.28) | Reference | 1.06 (0.91-1.24) | 1.43 (0.86-2.40) | .30 | .31 |
| **Cases diagnosed >9 years after enrollment (n = 634 pairs, case control matching 1:1)** | | | | | | | |
| 8-oxo-dG | | | | | | | |
| Age adjusted | 1.22 (0.96-1.54) | 1.08 (0.99-1.18) | Reference | 0.95 (0.90-1.01) | 0.92 (0.77-1.08) | .16 | .20 |
| Multivariable adjusted [c] | 1.17 (0.91-1.50) | 1.06 (0.97-1.17) | Reference | 0.96 (0.91-1.02) | 0.92 (0.78-1.09) | .31 | .34 |
| 8-oxo-Guo | | | | | | | |
| Age adjusted | 1.09 (0.84-1.43) | 1.04 (0.96-1.13) | Reference | 0.94 (0.87-1.01) | 0.80 (0.61-1.05) | .20 | .98 |
| Multivariable adjusted [c] | 1.02 (0.77-1.34) | 1.02 (0.94-1.12) | Reference | 0.93 (0.86-1.00) | 0.77 (0.58-1.02) | .18 | .55 |

Abbreviations: 8-oxo-dG, 8-Oxo-7,8-dihydro-2'-deoxyguanosine; 8-oxo-Guo, 7,8-dihydro-8-oxo-guanosine; SWHS, Shanghai Women's Health Study; CI, confidence interval; Cr, creatinine; OR, odds ratio.

[a] Case-control pairs were categorized into three subgroups based on the time interval between urine collection at baseline and cancer diagnosis during follow-up in the cases of the case-control pairs. P for heterogeneity across three time-interval groups (< 5 years, 5-9 years, and > 9 years) was 0.03 for 8-oxo-dG, and 0.07 for 8-oxo-Guo in the multivariable adjusted model.

[b] Cutoffs were based on the percentile distribution of 8-oxo-dG and 8-oxo-Guo levels in controls—for 8-oxo-dG (ng/mg Cr): 4.51 (10th), 6.57 (30th), 8.60 (50th), 11.80 (70th), and 18.17 (90th); and for 8-oxo-Guo (ng/mg Cr): 6.56 (10th), 9.54 (30th), 12.21 (50th), 16.19 (70th), and 25.56 (90th). The OR was estimated by using a conditional logistic regression model with restricted cubic-spline functions, with the 50th percentile of 8-oxo-dG and 8-oxo-Guo levels in controls treated as the reference.

[c] Multivariable adjusted for age at baseline, education, cigarette smoking, alcohol consumption, BMI, physical activity, menopausal status, regular use of vitamin supplements, regular use of aspirin and other nonsteroidal anti-inflammatory drugs, Charlson comorbidity score, family history of colorectal cancer in first-degree relatives, and total energy intake.

**Supplementary Table 8. Associations between baseline urinary levels of 8-oxo-dG and 8-oxo-Guo and subsequent risk of colorectal cancer by time interval between urine sample collection at baseline and cancer diagnosis during follow-up in SMHS [a]**

| | OR (95% CI) for colorectal cancer risk relative to reference, by percentile distribution of 8-oxo-dG and 8-oxo-Guo [b] | | | | | *P* for overall association | *P* for nonlinear association |
|---|---|---|---|---|---|---|---|
| | 10th | 30th | 50th | 70th | 90th | | |
| **Cases diagnosed <5 years after enrollment (n = 244 pairs, case control matching 1:1)** | | | | | | | |
| 8-oxo-dG | | | | | | | |
| Age adjusted | 2.06 (1.45-2.93) | 1.42 (1.20-1.69) | Reference | 0.63 (0.51-0.78) | 0.44 (0.31-0.62) | <.001 | .004 |
| Multivariable adjusted [c] | 2.14 (1.46-3.14) | 1.45 (1.21-1.74) | Reference | 0.62 (0.49-0.77) | 0.43 (0.30-0.62) | <.001 | .005 |
| 8-oxo-Guo | | | | | | | |
| Age adjusted | 1.56 (1.12-2.17) | 1.23 (1.06-1.44) | Reference | 0.75 (0.60-0.92) | 0.50 (0.32-0.78) | .008 | .06 |
| Multivariable adjusted [c] | 1.57 (1.10-2.23) | 1.24 (1.05-1.46) | Reference | 0.74 (0.59-0.93) | 0.45 (0.28-0.73) | .002 | .22 |
| **Cases diagnosed 5-9 years after enrollment (n = 324 pairs, case control matching 1:1)** | | | | | | | |
| 8-oxo-dG | | | | | | | |
| Age adjusted | 1.18 (0.91-1.54) | 1.09 (0.96-1.24) | Reference | 0.90 (0.76-1.05) | 0.81 (0.61-1.06) | .28 | .34 |
| Multivariable adjusted [c] | 1.05 (0.79-1.41) | 1.02 (0.89-1.18) | Reference | 0.96 (0.80-1.15) | 0.90 (0.67-1.22) | .56 | .95 |
| 8-oxo-Guo | | | | | | | |
| Age adjusted | 1.02 (0.76-1.37) | 1.01 (0.88-1.16) | Reference | 0.98 (0.82-1.17) | 0.92 (0.67-1.25) | .53 | .88 |
| Multivariable adjusted [c] | 0.97 (0.71-1.34) | 0.99 (0.85-1.15) | Reference | 1.01 (0.84-1.23) | 0.97 (0.69-1.35) | .58 | .66 |
| **Cases diagnosed >9 years after enrollment (n = 335 pairs, case control matching 1:1)** | | | | | | | |
| 8-oxo-dG | | | | | | | |
| Age adjusted | 0.73 (0.51-1.04) | 0.87 (0.74-1.02) | Reference | 1.10 (0.96-1.26) | 1.04 (0.75-1.44) | .21 | .09 |
| Multivariable adjusted [c] | 0.69 (0.47-1.00) | 0.85 (0.72-1.00) | Reference | 1.12 (0.97-1.29) | 1.06 (0.75-1.48) | .15 | .06 |
| 8-oxo-Guo | | | | | | | |
| Age adjusted | 0.74 (0.49-1.13) | 0.88 (0.74-1.05) | Reference | 1.11 (0.96-1.29) | 1.15 (0.81-1.63) | .34 | .25 |
| Multivariable adjusted [c] | 0.70 (0.45-1.08) | 0.86 (0.71-1.03) | Reference | 1.14 (0.98-1.32) | 1.18 (0.82-1.69) | .23 | .18 |

Abbreviations: 8-oxo-dG, 8-Oxo-7,8-dihydro-2'-deoxyguanosine; 8-oxo-Guo, 7,8-dihydro-8-oxo-guanosine; SMHS, Shanghai Men's Health Study; CI, confidence interval; Cr, creatinine; OR, odds ratio.

[a] Case-control pairs were categorized into three subgroups based on the time interval between urine collection at baseline and cancer diagnosis during follow-up in the cases of the case-control pairs. P for heterogeneity across three time-interval groups (< 5 years, 5-9 years, and > 9 years) was <0.001 for 8-oxo-dG, and 0.04 for 8-oxo-Guo in the multivariable adjusted model.

[b] Cutoffs were based on the percentile distribution of 8-oxo-dG and 8-oxo-Guo levels in controls—for 8-oxo-dG (ng/mg Cr): 4.51 (10th), 6.57 (30th), 8.60 (50th), 11.80 (70th), and 18.17 (90th); and for 8-oxo-Guo (ng/mg Cr): 6.56 (10th), 9.54 (30th), 12.21 (50th), 16.19 (70th), and 25.56 (90th). The OR was estimated by using a conditional logistic regression model with restricted cubic-spline functions, with the 50th percentile of 8-oxo-dG and 8-oxo-Guo levels in controls treated as the reference.

[c] Multivariable adjusted for age at baseline, education, cigarette smoking, alcohol consumption, BMI, physical activity, regular use of vitamin supplements, regular use of aspirin and other nonsteroidal anti-inflammatory drugs, Charlson comorbidity score, family history of colorectal cancer in first-degree relatives, and total energy intake.

**Supplementary Table 9. Associations between baseline urinary levels of 8-oxo-dG and 8-oxo-Guo and subsequent risk of colorectal cancer in SWHS and SMHS, stratified by cancer anatomical sites and by time interval between urine sample collection at baseline and cancer diagnosis during follow-up**

| By cancer anatomical sites (colon vs. rectal), and by time interval | OR (95% CI) for colorectal cancer risk relative to reference, by percentile distribution of 8-oxo-dG and 8-oxo-Guo [a,b] | | | | | *P* for overall association | *P* for nonlinear association | *P* for heterogeneity |
|---|---|---|---|---|---|---|---|---|
| | 10th | 30th | 50th | 70th | 90th | | | |
| **All participants (n = 1938 pairs, case control matching 1:1)** | | | | | | | | |
| 8-oxo-dG | | | | | | | | |
| Colon cancer (n = 1228 pairs) | 1.23 (1.03-1.46) | 1.10 (1.02-1.19) | Reference | 0.87 (0.81-0.93) | 0.68 (0.58-0.80) | <.001 | .63 | .006 |
| Rectal cancer (n = 710 pairs) | 1.15 (0.92-1.44) | 1.07 (0.96-1.18) | Reference | 0.95 (0.87-1.03) | 0.93 (0.81-1.07) | .43 | .27 | |
| 8-oxo-Guo | | | | | | | | |
| Colon cancer (n = 1228 pairs) | 1.16 (0.96-1.41) | 1.07 (0.99-1.16) | Reference | 0.92 (0.86-0.99) | 0.81 (0.70-0.94) | .02 | .50 | .24 |
| Rectal cancer (n = 710 pairs) | 0.97 (0.75-1.27) | 0.99 (0.89-1.11) | Reference | 1.00 (0.91-1.10) | 0.97 (0.81-1.16) | .85 | .73 | |
| **Colon cancer (n = 1228 pairs, case control matching 1:1)** | | | | | | | | |
| 8-oxo-dG | | | | | | | | |
| Cases diagnosed <5 years (n = 251 pairs) | 1.78 (1.21-2.63) | 1.31 (1.10-1.57) | Reference | 0.72 (0.60-0.86) | 0.51 (0.36-0.73) | <.001 | .09 | |
| Cases diagnosed 5-9 years (n = 335 pairs) | 1.23 (0.87-1.74) | 1.10 (0.94-1.30) | Reference | 0.89 (0.76-1.04) | 0.79 (0.60-1.04) | .24 | .48 | .12 |
| Cases diagnosed >9 years (n = 642 pairs) | 1.03 (0.80-1.32) | 1.03 (0.93-1.14) | Reference | 0.89 (0.82-0.97) | 0.66 (0.48-0.89) | .03 | .31 | |
| 8-oxo-Guo | | | | | | | | |
| Cases diagnosed <5 years (n = 251 pairs) | 1.28 (0.86-1.90) | 1.12 (0.94-1.34) | Reference | 0.86 (0.71-1.05) | 0.67 (0.45-0.99) | .14 | .66 | |
| Cases diagnosed 5-9 years (n = 335 pairs) | 1.16 (0.81-1.67) | 1.07 (0.91-1.26) | Reference | 0.93 (0.79-1.10) | 0.89 (0.67-1.19) | .70 | .47 | .72 |
| Cases diagnosed >9 years (n = 642 pairs) | 1.11 (0.84-1.48) | 1.05 (0.95-1.17) | Reference | 0.92 (0.85-1.00) | 0.76 (0.58-0.99) | .11 | .95 | |
| **Rectal cancer (n = 710 pairs, case control matching 1:1)** | | | | | | | | |
| 8-oxo-dG | | | | | | | | |
| Cases diagnosed <5 years (n = 178 pairs) | 1.88 (1.13-3.15) | 1.35 (1.07-1.71) | Reference | 0.69 (0.55-0.87) | 0.45 (0.30-0.69) | <.001 | .21 | .002 |

| | | | | | | | | |
|---|---|---|---|---|---|---|---|---|
| Cases diagnosed 5-9 years (n = 205 pairs) | 0.86 (0.56-1.32) | 0.93 (0.76-1.14) | Reference | 1.06 (0.87-1.29) | 1.03 (0.78-1.38) | .69 | .43 | |
| Cases diagnosed >9 years (n = 327 pairs) | 1.01 (0.71-1.44) | 0.99 (0.86-1.15) | Reference | 1.06 (0.94-1.20) | 1.27 (0.84-1.93) | .52 | .58 | |
| 8-oxo-Guo | | | | | | | | |
| Cases diagnosed <5 years (n = 178 pairs) | 1.61 (0.95-2.73) | 1.24 (0.98-1.58) | Reference | 0.76 (0.59-0.98) | 0.50 (0.31-0.81) | .02 | .38 | |
| Cases diagnosed 5-9 years (n = 205 pairs) | 1.01 (0.62-1.67) | 1.01 (0.81-1.26) | Reference | 1.00 (0.79-1.25) | 1.01 (0.69-1.47) | .99 | .93 | .02 |
| Cases diagnosed >9 years (n = 327 pairs) | 0.69 (0.45-1.07) | 0.87 (0.74-1.02) | Reference | 1.10 (0.98-1.23) | 1.19 (0.80-1.76) | .18 | .21 | |

Abbreviations: 8-oxo-dG, 8-Oxo-7,8-dihydro-2'-deoxyguanosine; 8-oxo-Guo, 7,8-dihydro-8-oxo-guanosine; SWHS, Shanghai Women's Health Study; SMHS, Shanghai Men's Health Study; CI, confidence interval; Cr, creatinine; OR, odds ratio.

[a] Cutoffs were based on the percentile distribution of 8-oxo-dG and 8-oxo-Guo levels in controls—for 8-oxo-dG (ng/mg Cr): 4.51 (10th), 6.57 (30th), 8.60 (50th), 11.80 (70th), and 18.17 (90th); and for 8-oxo-Guo (ng/mg Cr): 6.56 (10th), 9.54 (30th), 12.21 (50th), 16.19 (70th), and 25.56 (90th). The OR was estimated by using a conditional logistic regression model with restricted cubic-spline functions, with the 50th percentile of 8-oxo-dG and 8-oxo-Guo levels in controls treated as the reference.

[b] Multivariable adjusted for age at baseline, education, cigarette smoking, alcohol consumption, BMI, physical activity, regular use of vitamin supplements, regular use of aspirin and other nonsteroidal anti-inflammatory drugs, Charlson comorbidity score, family history of colorectal cancer in first-degree relatives, and total energy intake.

**Supplementary Table 10. Associations between baseline urinary levels of 8-oxo-dG and 8-oxo-Guo and subsequent risk of colorectal cancer in SWHS and SMHS, stratified by cancer stages and by time interval between urine sample collection at baseline and cancer diagnosis during follow-up [a]**

| By cancer stages (early vs. late), and by time interval | OR (95% CI) for colorectal cancer risk relative to reference, by percentile distribution of 8-oxo-dG and 8-oxo-Guo [b,c] | | | | | *P* for overall association | *P* for nonlinear association | *P* for heterogeneity |
|---|---|---|---|---|---|---|---|---|
| | 10th | 30th | 50th | 70th | 90th | | | |
| **All cases with known clinical factors (n = 1366 pairs, case control matching 1:1)** | | | | | | | | |
| 8-oxo-dG | | | | | | | | |
| Stage I and II (n=720 pairs) | 1.19 (0.94-1.51) | 1.09 (0.98-1.21) | Reference | 0.88 (0.81-0.97) | 0.71 (0.58-0.87) | .004 | .78 | .36 |
| Stage III and IV (n=646 pairs) | 1.43 (1.13-1.82) | 1.18 (1.06-1.31) | Reference | 0.86 (0.78-0.94) | 0.78 (0.66-0.92) | .003 | .02 | |
| 8-oxo-Guo | | | | | | | | |
| Stage I and II (n=720 pairs) | 0.96 (0.74-1.23) | 0.98 (0.88-1.10) | Reference | 1.00 (0.90-1.10) | 0.91 (0.74-1.10) | .35 | .44 | .31 |
| Stage III and IV (n=646 pairs) | 1.31 (0.99-1.74) | 1.13 (1.00-1.27) | Reference | 0.89 (0.80-0.98) | 0.79 (0.64-0.96) | .05 | .19 | |
| **Cases diagnosed <5 years after enrollment (n = 374 pairs, case control matching 1:1)** | | | | | | | | |
| 8-oxo-dG | | | | | | | | |
| Stage I and II (n=194 pairs) | 1.76 (1.11-2.79) | 1.31 (1.06-1.62) | Reference | 0.71 (0.58-0.88) | 0.48 (0.32-0.72) | .001 | .23 | .96 |
| Stage III and IV (n=180 pairs) | 2.24 (1.36-3.67) | 1.45 (1.16-1.81) | Reference | 0.67 (0.54-0.82) | 0.47 (0.30-0.74) | <.001 | .04 | |
| 8-oxo-Guo | | | | | | | | |
| Stage I and II (n=194 pairs) | 1.13 (0.69-1.84) | 1.06 (0.85-1.32) | Reference | 0.91 (0.70-1.17) | 0.68 (0.43-1.08) | .10 | .81 | .06 |
| Stage III and IV (n=180 pairs) | 3.27 (1.73-6.19) | 1.69 (1.28-2.24) | Reference | 0.58 (0.44-0.77) | 0.42 (0.23-0.77) | <.001 | .003 | |
| **Cases diagnosed 5-9 years after enrollment (n = 419 pairs, case control matching 1:1)** | | | | | | | | |
| 8-oxo-dG | | | | | | | | |
| Stage I and II (n=225 pairs) | 0.88 (0.55-1.43) | 0.95 (0.76-1.19) | Reference | 1.00 (0.80-1.24) | 0.75 (0.51-1.11) | .16 | .27 | .13 |
| Stage III and IV (n=194 pairs) | 1.49 (1.00-2.22) | 1.21 (1.00-1.46) | Reference | 0.82 (0.66-1.00) | 0.74 (0.53-1.03) | .15 | .06 | |
| 8-oxo-Guo | | | | | | | | |
| Stage I and II (n=225 pairs) | 0.73 (0.45-1.18) | 0.87 (0.70-1.08) | Reference | 1.15 (0.92-1.45) | 1.17 (0.78-1.76) | .42 | .19 | .11 |

| | | | | | | | | |
|---|---|---|---|---|---|---|---|---|
| Stage III and IV (n=194 pairs) | 1.69 (0.97-2.93) | 1.26 (0.99-1.62) | Reference | 0.78 (0.60-1.01) | 0.67 (0.44-1.01) | .15 | .09 | |
| **Cases diagnosed >9 years after enrollment (n = 573 pairs, case control matching 1:1)** | | | | | | | | |
| 8-oxo-dG | | | | | | | | |
| Stage I and II (n=301 pairs) | 1.07 (0.74-1.54) | 1.03 (0.89-1.20) | Reference | 0.96 (0.86-1.07) | 0.89 (0.68-1.17) | .70 | .93 | .80 |
| Stage III and IV (n=272 pairs) | 1.11 (0.73-1.67) | 1.05 (0.90-1.23) | Reference | 0.91 (0.80-1.04) | 0.75 (0.50-1.13) | .36 | .92 | |
| 8-oxo-Guo | | | | | | | | |
| Stage I and II (n=301 pairs) | 1.12 (0.73-1.71) | 1.04 (0.89-1.22) | Reference | 0.97 (0.86-1.08) | 0.94 (0.69-1.28) | .84 | .69 | .20 |
| Stage III and IV (n=272 pairs) | 0.79 (0.50-1.26) | 0.94 (0.80-1.11) | Reference | 0.96 (0.83-1.10) | 0.72 (0.44-1.20) | .26 | .15 | |

Abbreviations: 8-oxo-dG, 8-Oxo-7,8-dihydro-2'-deoxyguanosine; 8-oxo-Guo, 7,8-dihydro-8-oxo-guanosine; SWHS, Shanghai Women's Health Study; SMHS, Shanghai Men's Health Study; CI, confidence interval; Cr, creatinine; OR, odds ratio.

[a] Analyses were restricted to cases with known clinical stage and their individually matched controls (n = 1366 pairs).

[b] Cutoffs were based on the percentile distribution of 8-oxo-dG and 8-oxo-Guo levels in controls—for 8-oxo-dG (ng/mg Cr): 4.51 (10th), 6.57 (30th), 8.60 (50th), 11.80 (70th), and 18.17 (90th); and for 8-oxo-Guo (ng/mg Cr): 6.56 (10th), 9.54 (30th), 12.21 (50th), 16.19 (70th), and 25.56 (90th). The OR was estimated by using a conditional logistic regression model with restricted cubic-spline functions, with the 50th percentile of 8-oxo-dG and 8-oxo-Guo levels in controls treated as the reference.

[c] Multivariable adjusted for age at baseline, education, cigarette smoking, alcohol consumption, BMI, physical activity, regular use of vitamin supplements, regular use of aspirin and other nonsteroidal anti-inflammatory drugs, Charlson comorbidity score, family history of colorectal cancer in first-degree relatives, and total energy intake.

**Supplementary Table 11. Selected functional SNPs in DNA repair genes**

| Gene | SNP (rsID) | Chr. | Position (bp, GRCh38) | Alleles | Genomic Context |
|---|---|---|---|---|---|
| *EXO1* | rs3754093 | 1 | 241846814 | G>A | 5′ flanking region |
| | rs10802996 | 1 | 241847325 | G>C | 5′ flanking region |
| | rs1635517 | 1 | 241848731 | G>A | 5' UTR |
| | rs1776177 | 1 | 241848847 | C>T | 5' UTR |
| | rs735943 | 1 | 241866849 | A>G | His354Arg |
| | rs1047840 | 1 | 241878999 | A>G | Lys589Glu |
| | rs1776148 | 1 | 241879243 | A>G | Glu670Gly |
| | rs1635498 | 1 | 241881973 | C>T | Arg723Cys |
| | rs9350 | 1 | 241885372 | T>C | Leu756Pro |
| | rs851797 | 1 | 241889740 | G>A | 3' UTR |
| *MUTYH* | rs3219489 | 1 | 45331833 | G>C | His324Gln |
| *NUDT15* | rs186364861 | 13 | 48037798 | A>G | Ile18Val |
| | rs116855232 | 13 | 48045719 | T>C | Cys139Arg |
| *APEX1* | rs1760944 | 14 | 20454990 | G>T | 5′ flanking region |
| | rs2307486 | 14 | 20456045 | G>A | Val64Ile |
| | rs3136817 | 14 | 20456275 | C>T | Intron |
| | rs1130409 | 14 | 20456995 | G>T | Glu148Asp |
| *LIG3* | rs3744356 | 17 | 34986110 | T>C | Gly224Arg |
| | rs3135967 | 17 | 34986710 | G>A | Intron |
| | rs3135983 | 17 | 34991293 | T>G | Intron |
| | rs4796030 | 17 | 35003131 | C>A | Intron |
| | rs1052536 | 17 | 35004556 | T>C | Intron |
| | rs3744357 | 17 | 35007093 | T>C | Thr948= |
| *LIG1* | rs20581 | 19 | 48119170 | A>G | Asp802= |
| | rs10500298 | 19 | 48145214 | T>C | Intron |
| | rs20580 | 19 | 48151296 | G>T | Ala170= |
| | rs20579 | 19 | 48165573 | A>G | Non-coding transcript |
| | rs439132 | 19 | 48165657 | C>T | Intron |
| *XPC* | rs2228001 | 3 | 14145949 | G>T | Gln939Lys |
| | rs2228000 | 3 | 14158387 | A>G | Val499Ala |
| | rs1870134 | 3 | 14178523 | C>G | Val16Leu |
| | rs2607775 | 3 | 14178595 | G>C | Non-coding transcript |
| *OGG1* | rs1052133 | 3 | 9757089 | C>G | Ser326Cys |
| *NUDT1* | rs4866 | 7 | 2249951 | A>G | Met83Val |
| | rs1799832 | 7 | 2250887 | T>C | Asp119= |

Abbreviations: Chr., chromosome; SNP, single nucleotide polymorphism; rsID, reference SNP ID number; UTR, untranslated region.

**Supplementary Table 12. Associations of selected functional SNPs in DNA repair genes with urinary levels of 8-oxo-dG and 8-oxo-Guo among controls from SWHS and SMHS [a]**

| Gene | SNP (rsID) | Genotype [b] | EAF (%) | 8-oxo-dG [c] | | | 8-oxo-Guo [c] | | |
|---|---|---|---|---|---|---|---|---|---|
| | | | | Fold change (95% CI) [d] | *P* value | FDR-adjusted *P* value [e] | Fold change (95% CI) [d] | *P* value | FDR-adjusted *P* value [e] |
| *EXO1* | rs3754093 | G/A | 26.8 | 1.01 (0.97-1.05) | .53 | .84 | 0.99 (0.95-1.04) | .81 | .95 |
| | rs10802996 | G/C | 20.5 | 1.00 (0.96-1.05) | .99 | .99 | 1.01 (0.96-1.06) | .76 | .95 |
| | rs1635517 | G/A | 22.3 | 0.98 (0.94-1.02) | .27 | .76 | 0.98 (0.94-1.03) | .52 | .95 |
| | rs1776177 | C/T | 31.3 | 1.00 (0.97-1.04) | .83 | .98 | 0.99 (0.95-1.04) | .79 | .95 |
| | rs735943 | A/G | 22.5 | 0.98 (0.94-1.02) | .30 | .76 | 0.98 (0.93-1.03) | .40 | .95 |
| | rs1047840 | A/G | 19.1 | 1.03 (0.98-1.08) | .22 | .76 | 1.05 (0.99-1.11) | .09 | .95 |
| | rs1776148 | A/G | 16.8 | 1.02 (0.98-1.07) | .36 | .78 | 1.00 (0.95-1.06) | .98 | .98 |
| | rs1635498 | C/T | 14.0 | 0.96 (0.92-1.01) | .15 | .74 | 1.02 (0.96-1.08) | .47 | .95 |
| | rs9350 | T/C | 43.3 | 1.02 (0.98-1.06) | .29 | .76 | 1.01 (0.96-1.05) | .76 | .95 |
| | rs851797 | G/A | 42.6 | 1.00 (0.96-1.03) | .83 | .98 | 0.98 (0.94-1.02) | .30 | .95 |
| *MUTYH* | rs3219489 | G/C | 37.8 | 1.01 (0.98-1.05) | .46 | .84 | 1.01 (0.97-1.06) | .60 | .95 |
| *NUDT15* | rs186364861 | A/G | 0.6 | 0.99 (0.79-1.24) | .94 | .98 | 1.02 (0.78-1.33) | .91 | .95 |
| | rs116855232 | T/C | 12.1 | 1.06 (1.00-1.12) | **.04** | .39 | 1.00 (0.94-1.07) | .88 | .95 |
| *APEX1* | rs1760944 | G/T | 42.2 | 1.01 (0.97-1.04) | .70 | .98 | 1.02 (0.98-1.06) | .40 | .95 |
| | rs2307486 | G/A | 3.3 | 1.02 (0.92-1.12) | .74 | .98 | 1.03 (0.91-1.16) | .64 | .95 |
| | rs3136817 | C/T | 8.2 | 1.02 (0.96-1.09) | .47 | .84 | 0.99 (0.91-1.06) | .70 | .95 |
| | rs1130409 | G/T | 38.5 | 1.00 (0.97-1.04) | .92 | .98 | 1.00 (0.96-1.04) | .93 | .95 |
| *LIG3* | rs3744356 | T/C | 1.2 | 0.92 (0.78-1.08) | .32 | .76 | 0.93 (0.76-1.13) | .44 | .95 |
| | rs3135967 | G/A | 28.4 | 1.01 (0.97-1.05) | .72 | .98 | 1.00 (0.96-1.05) | .84 | .95 |
| | rs3135983 | T/G | 14.6 | 0.95 (0.91-1.00) | .07 | .39 | 0.96 (0.91-1.02) | .24 | .95 |
| | rs4796030 | C/A | 42.9 | 0.98 (0.95-1.02) | .32 | .76 | 0.99 (0.95-1.03) | .56 | .95 |
| | rs1052536 | T/C | 27.9 | 1.00 (0.96-1.04) | .88 | .98 | 1.00 (0.95-1.04) | .83 | .95 |
| | rs3744357 | T/C | 14.3 | 0.97 (0.92-1.02) | .19 | .75 | 0.97 (0.92-1.03) | .36 | .95 |
| *LIG1* | rs20581 | A/G | 13.5 | 0.94 (0.90-1.00) | **.03** | .39 | 0.96 (0.90-1.02) | .18 | .95 |
| | rs10500298 | T/C | 42.9 | 0.99 (0.95-1.02) | .50 | .84 | 1.01 (0.97-1.06) | .58 | .95 |
| | rs20580 | G/T | 31.0 | 0.96 (0.93-1.00) | .06 | .39 | 0.99 (0.95-1.04) | .70 | .95 |
| | rs20579 | A/G | 12.0 | 1.05 (1.00-1.11) | .07 | .39 | 1.02 (0.96-1.09) | .55 | .95 |
| | rs439132 | C/T | 16.7 | 1.00 (0.95-1.04) | .84 | .98 | 1.02 (0.97-1.08) | .46 | .95 |
| *XPC* | rs2228001 | G/T | 35.2 | 0.99 (0.95-1.02) | .52 | .84 | 1.01 (0.97-1.06) | .60 | .95 |
| | rs2228000 | A/G | 32.8 | 1.00 (0.96-1.04) | .95 | .98 | 0.99 (0.95-1.04) | .69 | .95 |
| | rs1870134 | C/G | 27.2 | 1.00 (0.97-1.04) | .82 | .98 | 1.00 (0.95-1.05) | .93 | .95 |
| | rs2607775 | G/C | 4.3 | 1.03 (0.94-1.12) | .51 | .84 | 1.01 (0.91-1.12) | .89 | .95 |
| *OGG1* | rs1052133 | C/G | 40.1 | 0.96 (0.93-0.99) | **.03** | .39 | 0.97 (0.93-1.02) | .21 | .95 |
| *NUDT1* | rs4866 | A/G | 3.1 | 0.93 (0.84-1.03) | .17 | .75 | 0.87 (0.77-0.98) | **.02** | .67 |
| | rs1799832 | T/C | 7.3 | 1.01 (0.94-1.08) | .82 | .98 | 1.08 (0.99-1.17) | .08 | .95 |

Abbreviations: 8-oxo-dG, 8-Oxo-7,8-dihydro-2'-deoxyguanosine; 8-oxo-Guo, 7,8-dihydro-8-oxo-guanosine; SWHS, Shanghai Women's Health Study; SMHS, Shanghai Men's Health Study; EAF, effect allele frequency; CI, confidence interval; SNP, single nucleotide polymorphism; rsID, reference SNP ID number.

[a] Analyses were restricted to the controls with data available on genetic variants (n = 1711).

[b] Effect allele/other allele.

[c] Covariates included in the liner regression model were age, education, cigarette smoking, alcohol consumption, BMI, physical activity, sex, regular use of vitamin supplements, regular use of aspirin and other nonsteroidal anti-inflammatory drugs, Charlson comorbidity score, and total energy intake.

[d] Estimated fold change (exponential function of regression coefficient) of the urinary levels of 8-oxo-dG and 8-oxo-Guo per one dosage increment in the effect allele for each genetic variant.

[e] FDR *p*-values were adjusted for multiple testing for selected 35 functional SNPs.

**Supplementary Table 13. Associations between selected functional SNPs in DNA repair genes and risk of colorectal cancer in SWHS and SMHS [a]**

| Gene | SNP (rsID) | Genotype [b] | EAF (%) | Effect estimate [c] | | FDR-adjusted *P* value [e] |
|---|---|---|---|---|---|---|
| | | | | OR (95% CI) [d] | *P* value | |
| *EXO1* | rs3754093 | G/A | 27.0 | 1.00 (0.89-1.12) | .95 | .99 |
| | rs10802996 | G/C | 21.2 | 1.08 (0.95-1.22) | .22 | .98 |
| | rs1635517 | G/A | 22.2 | 0.97 (0.86-1.10) | .65 | .99 |
| | rs1776177 | C/T | 31.6 | 1.02 (0.92-1.14) | .66 | .99 |
| | rs735943 | A/G | 22.4 | 0.97 (0.86-1.09) | .62 | .99 |
| | rs1047840 | A/G | 19.5 | 1.06 (0.93-1.21) | .36 | .99 |
| | rs1776148 | A/G | 17.0 | 1.03 (0.89-1.18) | .71 | .99 |
| | rs1635498 | C/T | 14.2 | 1.06 (0.91-1.22) | .47 | .99 |
| | rs9350 | T/C | 42.6 | 0.94 (0.85-1.04) | .25 | .98 |
| | rs851797 | G/A | 43.1 | 1.02 (0.92-1.13) | .68 | .99 |
| *MUTYH* | rs3219489 | G/C | 37.5 | 0.95 (0.86-1.06) | .36 | .99 |
| *NUDT15* | rs186364861 | A/G | 0.7 | 1.08 (0.59-1.98) | .81 | .99 |
| | rs116855232 | T/C | 12.3 | 1.05 (0.90-1.22) | .56 | .99 |
| *APEX1* | rs1760944 | G/T | 42.2 | 1.00 (0.90-1.11) | .99 | .99 |
| | rs2307486 | G/A | 3.4 | 1.01 (0.77-1.34) | .92 | .99 |
| | rs3136817 | C/T | 8.7 | 1.16 (0.97-1.40) | .11 | .64 |
| | rs1130409 | G/T | 39.3 | 1.07 (0.97-1.19) | .17 | .87 |
| *LIG3* | rs3744356 | T/C | 1.1 | 0.99 (0.60-1.63) | .96 | .99 |
| | rs3135967 | G/A | 28.5 | 1.00 (0.89-1.12) | .97 | .99 |
| | rs3135983 | T/G | 14.7 | 1.05 (0.91-1.21) | .51 | .99 |
| | rs4796030 | C/A | 43.1 | 1.01 (0.91-1.12) | .82 | .99 |
| | rs1052536 | T/C | 27.9 | 0.99 (0.88-1.11) | .80 | .99 |
| | rs3744357 | T/C | 14.5 | 1.06 (0.91-1.22) | .46 | .99 |
| *LIG1* | rs20581 | A/G | 13.9 | 1.05 (0.90-1.21) | .55 | .99 |
| | rs10500298 | T/C | 42.4 | 0.96 (0.86-1.06) | .39 | .99 |
| | rs20580 | G/T | 30.5 | 0.95 (0.85-1.06) | .39 | .99 |
| | rs20579 | A/G | 11.8 | 0.96 (0.82-1.12) | .62 | .99 |
| | rs439132 | C/T | 15.9 | 0.88 (0.76-1.02) | .08 | .64 |
| *XPC* | rs2228001 | G/T | 36.7 | 1.13 (1.02-1.25) | **.02** | .49 |
| | rs2228000 | A/G | 32.9 | 1.00 (0.90-1.12) | .95 | .99 |
| | rs1870134 | C/G | 26.0 | 0.89 (0.79-0.99) | **.04** | .49 |
| | rs2607775 | G/C | 3.9 | 0.80 (0.61-1.04) | .09 | .64 |
| *OGG1* | rs1052133 | C/G | 40.1 | 1.00 (0.90-1.11) | .98 | .99 |
| *NUDT1* | rs4866 | A/G | 3.5 | 1.33 (1.01-1.76) | **.04** | .49 |
| | rs1799832 | T/C | 7.4 | 1.03 (0.85-1.24) | .78 | .99 |

Abbreviations: SWHS, Shanghai Women's Health Study; SMHS, Shanghai Men's Health Study; EAF, effect allele frequency; CI, confidence interval; SNP, single nucleotide polymorphism; rsID, reference SNP ID number; OR, odds ratio.

[a] Analyses were restricted to case-control pairs with data available on genetic variants (n = 1545 pairs).

[b] Effect allele/other allele.

[c] Covariates included in the conditional logistic regression model were age at baseline, education, cigarette smoking, alcohol consumption, BMI, physical activity, regular use of vitamin supplements, regular use of aspirin and other nonsteroidal anti-inflammatory drugs, Charlson comorbidity score, family history of colorectal cancer in first-degree relatives, and total energy intake.

[d] OR for colorectal cancer risk per one dosage increment in the effect allele for each genetic variant.

[e] FDR p-values were adjusted for multiple testing for selected 35 functional SNPs.

**Supplementary Table 14. Associations between baseline urinary levels of 8-oxo-dG and 8-oxo-Guo and subsequent risk of colorectal cancer overall and by time interval between urine sample collection at baseline and cancer diagnosis during follow-up in SWHS and SMHS, after further adjustment for selected functional SNPs in DNA repair genes**

| | OR (95% CI) for colorectal cancer risk relative to reference, by percentile distribution of 8-oxo-dG and 8-oxo-Guo [a] | | | | | *P* for overall association | *P* for nonlinear association |
|---|---|---|---|---|---|---|---|
| | 10th | 30th | 50th | 70th | 90th | | |
| **All participants (n = 1545 pairs, case control matching 1:1)** | | | | | | | |
| 8-oxo-dG [b] | 1.30 (1.11, 1.52) | 1.13 (1.05, 1.21) | Reference | 0.89 (0.84, 0.95) | 0.83 (0.75, 0.93) | <.001 | .01 |
| 8-oxo-Guo [c] | 1.10 (0.92, 1.31) | 1.04 (0.97, 1.12) | Reference | 0.96 (0.90, 1.02) | 0.90 (0.80, 1.03) | .28 | .53 |
| **Cases diagnosed <5 years after enrollment (n = 424 pairs, case control matching 1:1)** | | | | | | | |
| 8-oxo-dG [b] | 1.90 (1.40, 2.57) | 1.35 (1.18, 1.55) | Reference | 0.70 (0.61, 0.80) | 0.49 (0.37, 0.64) | <.001 | .02 |
| 8-oxo-Guo [c] | 1.49 (1.09, 2.03) | 1.20 (1.04, 1.38) | Reference | 0.79 (0.68, 0.93) | 0.57 (0.42, 0.76) | .001 | .21 |
| **Cases diagnosed 5-9 years after enrollment (n = 511 pairs, case control matching 1:1)** | | | | | | | |
| 8-oxo-dG [b] | 1.11 (0.84, 1.46) | 1.05 (0.92, 1.19) | Reference | 0.95 (0.83, 1.07) | 0.90 (0.74, 1.09) | .56 | .60 |
| 8-oxo-Guo [c] | 1.02 (0.76, 1.38) | 1.01 (0.88, 1.15) | Reference | 0.99 (0.86, 1.14) | 1.01 (0.80, 1.27) | .95 | .84 |
| **Cases diagnosed >9 years after enrollment (n = 610 pairs, case control matching 1:1)** | | | | | | | |
| 8-oxo-dG [b] | 1.06 (0.81, 1.39) | 1.02 (0.92, 1.14) | Reference | 1.00 (0.93, 1.08) | 1.05 (0.88, 1.27) | .75 | .55 |
| 8-oxo-Guo [c] | 0.97 (0.71, 1.32) | 0.99 (0.88, 1.11) | Reference | 1.01 (0.93, 1.10) | 1.04 (0.79, 1.36) | .95 | .91 |

Abbreviations: 8-oxo-dG, 8-Oxo-7,8-dihydro-2'-deoxyguanosine; 8-oxo-Guo, 7,8-dihydro-8-oxo-guanosine; SWHS, Shanghai Women's Health Study; SMHS, Shanghai Men's Health Study; CI, confidence interval; Cr, creatinine; OR, odds ratio.

[a] Cutoffs were based on the percentile distribution of 8-oxo-dG and 8-oxo-Guo levels in controls—for 8-oxo-dG (ng/mg Cr): 4.51 (10th), 6.57 (30th), 8.60 (50th), 11.80 (70th), and 18.17 (90th); and for 8-oxo-Guo (ng/mg Cr): 6.56 (10th), 9.54 (30th), 12.21 (50th), 16.19 (70th), and 25.56 (90th). The OR was estimated by using a conditional logistic regression model with restricted cubic-spline functions, with the 50th percentile of 8-oxo-dG and 8-oxo-Guo levels in controls treated as the reference.

[b] Covariates in the multivariable model included age at baseline, education, cigarette smoking, alcohol consumption, BMI, physical activity, regular use of vitamin supplements, regular use of aspirin and other nonsteroidal anti-inflammatory drugs, Charlson comorbidity score, family history of colorectal

cancer in first-degree relatives, and total energy intake. Additionally, 6 functional SNPs (rs116855232, rs20581, rs1052133, rs2228001, rs1870134, and rs4866) that showed a nominally significant association with urinary concentrations of 8-oxo-dG (Supplementary Table 13) or colorectal cancer risk (Supplementary Table 14) were included.

[c] Covariates in the multivariable model included age at baseline, education, cigarette smoking, alcohol consumption, BMI, physical activity, regular use of vitamin supplements, regular use of aspirin and other nonsteroidal anti-inflammatory drugs, Charlson comorbidity score, family history of colorectal cancer in first-degree relatives, and total energy intake. Additionally, 3 functional SNPs (rs2228001, rs1870134, and rs4866) that showed a nominally significant association with urinary concentrations of 8-oxo-Guo (Supplementary Table 13) or colorectal cancer risk (Supplementary Table 14) were included.

15. Liu J, Zheng J, Guo Y, et al. Association between APE1 rs1760944 and

rs1130409 polymorphism with prostate cancer risk: A systematic review and meta-analysis. *Medicine (Baltimore).* 2021;100(46):e27630.
